# Exploring the role of nicotine and smoking in sleep behaviours: A multivariable Mendelian Randomisation study

**DOI:** 10.1101/2024.08.01.24311349

**Authors:** Stephanie Page, Mark Gibson, Marcus R. Munafò, Jasmine Khouja, Rebecca C. Richmond

**Affiliations:** MRC Integrative Epidemiology Unit, University of Bristol, Bristol, United Kingdom, BS8 2BN; Population Health Sciences, Bristol Medical School, University of Bristol, Bristol, United Kingdom, BS8 2BN; School of Psychological Sciences, University of Bristol, Bristol, United Kingdom, BS8 1TU

**Author notes:** Authors have contributed equally to the manuscript (Jasmine Khouja, Rebecca C. Richmond). Corresponding author: Stephanie Page, Address: Oakfield House, Oakfield Grove, Clifton, Bristol, BS8 2BN.

**Keywords:** Mendelian randomisation, genetics, smoking, nicotine, chronotype, sleep, napping

## Abstract

Research has shown bidirectional relationships between smoking and adverse sleep behaviours, including late chronotype and insomnia, but the underlying mechanisms are not understood. One potential driver is nicotine, but its role in sleep is unclear. For this study, we estimated the direct effect of nicotine on six sleep behaviours measured in UK Biobank (chronotype, ease of getting up in the morning, insomnia symptoms, napping, daytime sleepiness and sleep duration). We conducted a Mendelian randomisation (MR) study to explore whether nicotine metabolism has a causal effect on these sleep behaviours. We explored whether the effects could be explained by regular nicotine exposure using genetic proxies of the nicotine metabolite ratio (NMR) and cigarettes per day (CPD) in a multivariable MR design. We found a higher NMR (indicating lower levels of circulating nicotine per cigarette smoked) decreased the likelihood of being an evening person when accounting for CPD in current (β = −0.04, 95%CI −0.06 to −0.02, p < 0.001) and ever smokers (β = −0.03 95%CI −0.04 to −0.01, p = 0.003). A higher NMR also increased the ease of getting up (β = 0.02, 95%CI 0.01 to 0.04, p = 0.015) and likelihood of napping (β = 0.02, 95%CI CI 0.002 to 0.03, p = 0.029) in current smokers. Increased nicotine exposure may directly affect sleep and could underlie relationships between smoking and sleep behaviours identified previously. Sleep could also be impacted in individuals using nicotine delivery systems or using nicotine replacement therapies. Further research is warranted to strengthen this conclusion.

## Introduction

Prevalence estimates of insomnia vary depending on its definition, but approximately 10% of adults in Europe have persistent difficulty in initiating and maintaining sleep coupled with impairments in daily functioning (chronic insomnia) (Steiger et al., 2023). Sleep disturbances such as insomnia are associated with an increased risk of many burdensome physical health problems, including cardiovascular disease, type 2 diabetes, obesity, heart failure and all-cause mortality (Chaput et al., 2020; Jike et al., 2018; Knutson & von Schantz, 2018; Mahmood et al., 2021; Sofi et al., 2014). They have also been associated with multiple mental health disorders, including depression and anxiety (Hertenstein et al., 2019). Other sleep traits (e.g., evening chronotype - feeling more awake later in the day) have also been linked to physical and mental health issues such as hypertension, diabetes and mood disorders (Partonen, 2015). Therefore, identifying and targeting modifiable risk factors that cause sleep problems could improve mental and physical health as well as sleep.

Smoking has been previously linked with several sleep behaviours. In the UK Biobank study, current smoking is cross-sectionally associated with an increased likelihood of longer sleep duration (>9 hours/day) compared to never-smokers (odds ratio [OR] = 1.47, 95% confidence interval [CI] 1.17 to 1.85, p = 0.001) (Boakye et al., 2018). Further, individuals in UK Biobank who consider themselves an evening person are more likely to smoke (Patterson et al., 2016). In addition, smokers in UK Biobank are more likely to report insomnia, very short and short sleep vs normal sleep duration (and report even higher levels of these if they have been smoking at night) (Nuñez et al., 2021). Smokers in this cohort are also more likely to report daytime sleepiness than non-smokers (Costa & Esteves, 2018). However, the cross-sectional design of these previous studies means that the direction of these relationships is unclear.

A possible driver of a causal pathway between smoking and sleep is nicotine. As nicotine is a stimulant, it is possible that sleep disruption in smokers is a result of its stimulating effects (Saint-Mleux et al., 2004; Zhang et al., 2008). Smokers are also more likely than non-smokers to report daytime sleepiness because of sleep disturbances (Patterson et al., 2019). As defined by Khantzian (2003), individuals using substances may do so to alleviate discomfort (known as the self-medication hypothesis). Therefore, it is plausible that smokers also use the stimulating effects of nicotine to alleviate daytime sleepiness. Additionally, a meta-analysis of 52 studies has shown that nicotine withdrawal following smoking cessation may be exacerbated by insomnia and therefore increase the risk of relapse (Jaehne et al., 2009). However, the specific effect of nicotine cannot easily be separated from the effects of exposure to (or withdrawal from) other constituents of tobacco smoke in these studies (McNeill et al., 2022).

Nicotine replacement therapies (NRT) deliver nicotine without the harmful constituents of tobacco smoke. Their use has been associated with sleep disturbances in smokers trying to quit (Patterson et al., 2019). However, it is not possible to ascertain the effects of regular nicotine use on sleep patterns in these studies, since NRT tends to be used for relatively short periods (Shahab et al., 2018). The impact of regular nicotine use on sleep are important to understand, given the increase in people regularly using e-cigarettes, which can contain nicotine but far fewer harmful toxicants and chemicals than are found in cigarette smoke.

An approach which allows us to infer the causal nature and direction of observed relationships is Mendelian randomisation (MR). MR is a form of instrumental variable analysis in which genetic variants are used as a proxy measure for the modifiable exposure they are associated with to determine its relationship with an outcome of interest. If the core assumptions of MR are met, this approach allows us to estimate a causal effect (Davies et al., 2018). MR has been previously used to investigate the causal relationship between smoking and sleep (Gibson et al., 2018; Pasman et al., 2020). Using this approach, smoking heaviness has been found to decrease the likelihood of having a morning preference (Gibson et al., 2018), while smoking initiation has been found to increase the likelihood of insomnia (Pasman et al., 2020).

Given the methodological issues of disentangling the impact of nicotine from the impact of other chemicals and toxicants contained in cigarettes and e-cigarettes, MR could be a useful method to employ to explore the effects of nicotine on sleep. Khouja et al. (2021) have previously used an extension of MR, multivariable MR (MVMR; Sanderson et al., 2019) to differentiate between the effects of nicotine and the other constituents of tobacco smoke on health outcomes among people who smoke. The model used the nicotine metabolite ratio (NMR) as a proxy for nicotine exposure. NMR is a measure of how quickly a person metabolises nicotine, and therefore is an indicator of how much circulating nicotine a person will have given a fixed level of nicotine exposure (Sofuoglu et al., 2012). It is the ratio of two major nicotine metabolites: 3’hydroxycotinine/cotinine. Research suggests that the higher an individual’s NMR, the more cigarettes they are likely to smoke, as they require a higher dosage to feel the same effects as someone with a lower NMR (West et al., 2011).

In this study, we aimed to examine the direct effect of nicotine (NMR) on six self-reported sleep behaviours: chronotype, ease of getting up in the morning, insomnia symptoms, napping, daytime sleepiness and sleep duration. We did this while accounting for smoking heaviness, within an MVMR framework previously used by Khouja et al. (2021). This framework will provide insight into whether nicotine plays a part in the relationship between smoking and sleep behaviours.

## Methods

### Data sources

The data for these analyses were sourced from published Genome-wide association study (GWAS) summary-level data, or GWAS conducted within the UK Biobank (summary-level data derived from individual-level data accessed via a UK Biobank application; project 9142). Individual-level data refers to data collected on individual participants within a study, whereas summary-level statistics are aggregated from individual-level statistics to give the overall association for genetic variants of interest.

### NMR GWAS

Genetic variants (N SNPs = 6) conditionally independently associated with NMR (at the p-value threshold p = <5×10^-8^) were identified using summary-level data from a recent GWAS of NMR in 5,185 current smokers (Buchwald et al., 2021). These were drawn from five cohorts of European ancestry (three from Finland, one from North America and one from Australia). Details of each cohort are provided by the authors in the GWAS supplementary materials (Buchwald et al., 2021). Informed consent was given by participants in each of the original cohort studies. Detail of ethics approval for these studies is also given by the authors in the supplementary materials (Buchwald et al., 2021). Current smokers were defined as those with cotinine levels ≥10lng/ml. Anything below this was interpreted as non-daily smoking and these individuals were therefore excluded as NMR measurements were considered unstable. Details of the variant inclusion criteria and quality control of GWAS summary results are also available in the paper. Of the seven single nucleotide polymorphisms (SNPs) which Buchwald et al., 2021 identified as conditionally independent, one SNP (rs117090198) generated a p-value that was considerably higher than the genome-wide significant threshold (p = 0.978) prior to the conditional analysis and was omitted from our analyses to avoid introducing noise into the NMR instrument.

### Cigarettes per day GWAS

Fifty-five SNPs associated with CPD (at the p-value threshold p = <5×10^-8^) were used as a proxy measure of smoking heaviness based on publicly available data from the GWAS and Sequencing Consortium of Alcohol and Nicotine use (GSCAN) (Liu et al., 2019). For the purposes of our analyses, SNPs were selected for inclusion based on the p-value from the conditional analyses where the sample included participants from 23andMe. Liu et al. (2019) identified conditional SNPs using Conditional and joint association analysis (COJO) in the software Genome-wide Complex Trait Analysis (GCTA). The betas used for weighting in our analyses were derived in a GWAS excluding UK Biobank (to minimise sample overlap) and 23andMe (due to data sharing restrictions). CPD was defined as the average number of cigarettes smoked per day as a current or former smoker, measured in 337,334 individuals of European ancestry. Never smokers were excluded. Details of the quality control for summary statistics on each cohort are provided by the authors in their paper. This study received ethics approval from the University of Minnesota Institutional Review Board and all participants provided informed consent.

### Sleep trait GWAS

Summary-level data on six self-reported sleep behaviours (namely chronotype, ease of getting up in the morning, insomnia symptoms, napping, daytime sleepiness and sleep duration) were obtained from GWAS that were performed using UK Biobank data.

UK Biobank is a population-based health research resource consisting of approximately 500,000 people, aged between 38 years and 73 years, who were recruited between the years 2006 and 2010 from across the UK (Allen et al., 2014). Particularly focused on identifying determinants of human diseases in middle-aged and older individuals, participants provided a range of information (data available at www.ukbiobank.ac.uk). A full description of the study design, participants, and quality control (QC) methods have been described in detail previously (Collins, 2012). UK Biobank received ethics approval from the Research Ethics Committee (REC reference for UK Biobank is 11/NW/0382). Details of participant consent is given in the UK Biobank Ethics and Governance Framework (UK Biobank, 2023). This research has been conducted using the UK Biobank Resource under application number 9142.

Six self-reported sleep behaviours were measured: chronotype, ease of getting up in the morning, insomnia symptoms, napping, daytime sleepiness and sleep duration. All six measures were continuous or ordinal. Details on the sample size, questions and response codes for each are available in the Supplementary Methods.

Since the GWAS for NMR and CPD were conducted using data from smokers and former smokers (regarding their behaviour when they were a smoker), the SNPs identified are not applicable to non-smokers. Thus, we conducted GWAS for the sleep outcomes stratified by smoking status (‘current’, ‘ever, ‘former’ and ‘never’ smokers) using the MRC IEU UK Biobank GWAS pipeline.

This pipeline uses the software package BOLT-LMM (v2.3), accounting for relatedness and population stratification via a linear mixed model, as well as adjusting for age, sex and genotyping chip (Elsworth et al., 2019). Further details of this are available in the Supplementary Methods. We stratified smokers into these four groups: ever smokers, current smokers, former smokers, never smokers. Effects observed in ‘current smokers’ – those who smoked occasionally, most days or daily – are most relevant to our research question. We explored effects among ‘ever smokers’ (which encompassed ‘current’ and ‘former’ smokers) to investigate the effects with increased statistical power. Effects in ‘former smokers’ (not currently smoking but used to smoke occasionally, most days or daily) indicate whether effects could be recoverable by smoking cessation. Finally, effects observed in ‘never smokers’ (those who have tried smoking once or twice, or never) indicate pleiotropic effects or population stratification as these associations cannot be due to a causal impact of smoking (i.e., cannot have been caused by the individual smoking as they have never smoked).

### Statistical analysis

#### Univariable MR

We first used a two-sample univariable MR framework (specifically inverse variance weighted – IVW, MR-Egger, weighted median and weighted mode models) to investigate the total causal effect of each of our exposures (NMR and CPD) on sleep behaviours measured in UK Biobank. Data were harmonised to ensure that SNP-exposure and SNP-outcome effects corresponded to the same effect allele.

When conducting MR, three core assumptions must be met (Davies et al., 2018). These are specified in the Supplementary Methods. To check that the core assumptions of MR were met, we conducted sensitivity analyses. To check the ‘relevance’ assumption, we tested instrument strength using an F statistic for MR. An F statistic that exceeded the conventional threshold of 10 was taken to indicate good instrument strength (Sanderson et al., 2019). To assess the ‘exclusion restriction’ assumption, we tested for the presence of horizontal pleiotropy using MR-Egger regression, single SNP analysis and funnel plots (Supplementary Figures S1-S18). A non-zero intercept term in MR-Egger regression term is interpreted as the directional pleiotropic effect (Hemani et al., 2018). Therefore, an intercept other than zero was taken to indicate the presence of horizontal pleiotropy. Single SNP analysis was used to explore whether individual SNPs were associated with the outcome independent of exposure-outcome association (Burgess et al., 2017). Associations were therefore interpreted as the presence of horizontal pleiotropy. We used funnel plots to detect the presence of horizontal pleiotropy by visually inspecting for asymmetry in estimates (Bowden et al., 2015). We also assessed heterogeneity using Cochran’s Q statistic to check this assumption, where we interpreted a Cochran’s Q value that exceeded the degrees of freedom (N SNPs minus 1) as an indication of variation across effect estimates (Greco M et al., 2015). Cochran’s Q values exceeding the number of SNPs included in the instrument indicate heterogeneity. Further, leave-one-out analyses were conducted to assess whether the exposure-outcome association remained after individual SNPs were removed. In this sensitivity analysis, if the causal effect estimate attenuates towards the null when a single SNP is omitted, this suggests that the outlier may have biased the causal estimate (i.e., through horizontal pleiotropy). We were unable to directly assess the ‘independence’ assumption but since the included GWAS were restricted to individuals of European ancestry and/or included genetic principal components as covariates, this reduced potential confounding due to major population stratification.

Negative control analyses (i.e., analyses using outcome data from never smokers) were also used to indicate potential violation of the ‘independence’ and ‘exclusion restriction’ assumptions. It would not be possible for a genuine causal effect of smoking or nicotine to be observed in never smokers as they would have never smoked. Therefore, where analyses among never smokers show evidence of a causal effect on sleep behaviours, we assume that the effects are due to either pleiotropy or population stratification, which may bias our results among current, ever and former smokers.

### Multivariable MR

We used a two-sample MVMR framework to investigate the direct effect of NMR on sleep behaviours when accounting for CPD (and vice versa). An additional clumping stage is needed for MVMR to ensure independence of the exposure SNPs with each other, so data were clumped (LD r^2^<0.1, clumping window >500kb) and harmonised. The results reflect the effects of a per standard deviation increase in NMR on the outcome. When accounting for CPD in the MVMR model, a higher NMR was taken to reflect lower levels of circulating nicotine per cigarette smoked (as nicotine would have been cleared from the system).

The three core assumptions for MVMR are (Sanderson, 2021):

1. The genetic variants must be associated with each exposure of interest given all other exposures in the model (the ‘relevance’ assumption)
2. They must be independent of the outcome given all of the exposures (the ‘exclusion restriction’ assumption)
3. They must be independent of any confounders of the exposures and outcome (the ‘independence’ or ‘exchangeability’ assumption)

To test the assumptions of our MVMR models, we conducted sensitivity analyses. Firstly, to test the ‘relevance’ assumption, we assessed instrument strength using a conditional F statistic for MVMR, taking estimates over 10 to indicate good instrument strength (Sanderson et al., 2021). To assess the ‘exclusion restriction’ assumption, we tested for the presence of potential horizontal pleiotropy using Cochran’s Q (Sanderson et al., 2019). A statistic larger than the number of SNPs included in the instrument was interpreted as potential horizontal pleiotropy.

MR and MVMR analyses were conducted using the software package R (version 4. 0. 3). R packages used were TwoSampleMR (version 0. 5. 6) and MVMR (version 0.3). The MR analysis performed in this manuscript was done in accordance with the STROBE-MR guidelines and a checklist is disclosed in the Supplementary Methods (Skrivankova et al., 2021).

## Results

The numbers of UK Biobank individuals included in the GWAS of each sleep behaviour stratified by smoking status are shown in Table 1.

**Table 1.**
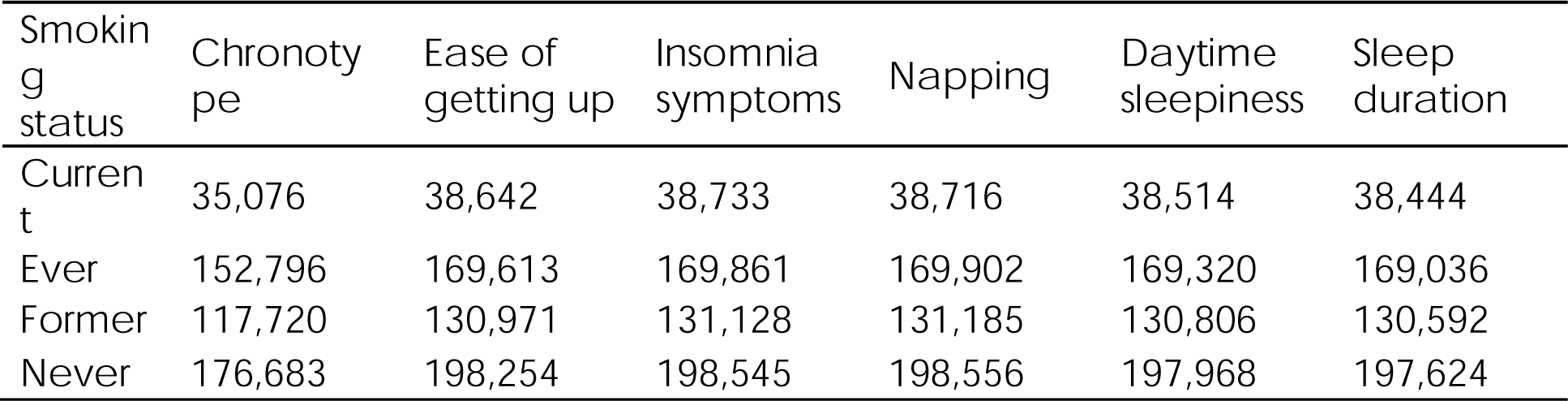
Number of UK Biobank participants with sleep behaviour data stratified by smoking status.

| Smoking status | Chronotype | Ease of getting up | Insomnia symptoms | Napping | Daytime sleepiness | Sleep duration |
| --- | --- | --- | --- | --- | --- | --- |
| Current | 35,076 | 38,642 | 38,733 | 38,716 | 38,514 | 38,444 |
| Ever | 152,796 | 169,613 | 169,861 | 169,902 | 169,320 | 169,036 |
| Former | 117,720 | 130,971 | 131,128 | 131,185 | 130,806 | 130,592 |
| Never | 176,683 | 198,254 | 198,545 | 198,556 | 197,968 | 197,624 |

### Univariable MR

#### Total effects of NMR and CPD on sleep outcomes

The F statistic for NMR and CPD were 450.99 and 130.22, suggesting good instrument strength in the univariable models. There was limited evidence for heterogeneity in the SNP estimates of NMR on the sleep outcomes (Supplementary Table S7), while more heterogeneity was observed for the CPD instrument across the sleep outcomes and smoking subgroups (Supplementary Table S8), indicative of possible horizontal pleiotropy. Results from a pleiotropy-robust method (MV-Egger analyses) are also presented.

Results from the UVMR-IVW analyses are presented in Supplementary Figure 19 and Supplementary Table S3 while UVMR-Egger analyses are presented in Supplementary Table S4.

Using MR-IVW, we found evidence that increased NMR decreased the likelihood of being an evening chronotype in current, ever and former smokers (i.e., not in never smokers); increased the ease of getting up among current and ever smokers; and increased napping and daytime sleepiness, while decreasing sleep duration among current smokers (Supplementary Table S3).

Meanwhile, increased CPD increased the likelihood of evening chronotype in current, ever and former smokers; decreased the ease of getting up among current and ever smokers; decreased daytime sleepiness in current and ever smokers, but increased daytime napping among current smokers; and increased sleep duration in ever and former smokers (Supplementary Table S3).

There was little evidence for an effect of either NMR on CPD on insomnia symptoms in the UVMR analysis (Supplementary Table S3).

There was some evidence that genetically-proxied CPD was associated with chronotype and ease of getting up among never smokers, and between NMR and daytime sleepiness among never smokers, indicating potential violation of the independence and/or exclusion restriction assumptions (Supplementary Table S3). However, findings from MR-Egger analysis were largely consistent with those in the IVW, albeit with slightly wider confidence intervals (Supplementary Table S4).

### Multivariable MR

In multivariable models, conditional F statistics for both NMR (F = 24.50) and CPD (F = 33.14) indicated good instrument strength (Sanderson et al., 2019). As the Cochran’s Q statistic was greater than the number of SNPs across all IVW-MVMR analyses (Supplementary Table 10), indicating heterogeneity, results from a pleiotropy-robust method (MVMR-Egger analyses) are also presented.

Results from the MVMR-IVW analyses are presented in Figure 1 and Supplementary Table S10 while MVMR-Egger analyses are presented in Supplementary Table S11.

**Figure 1.**
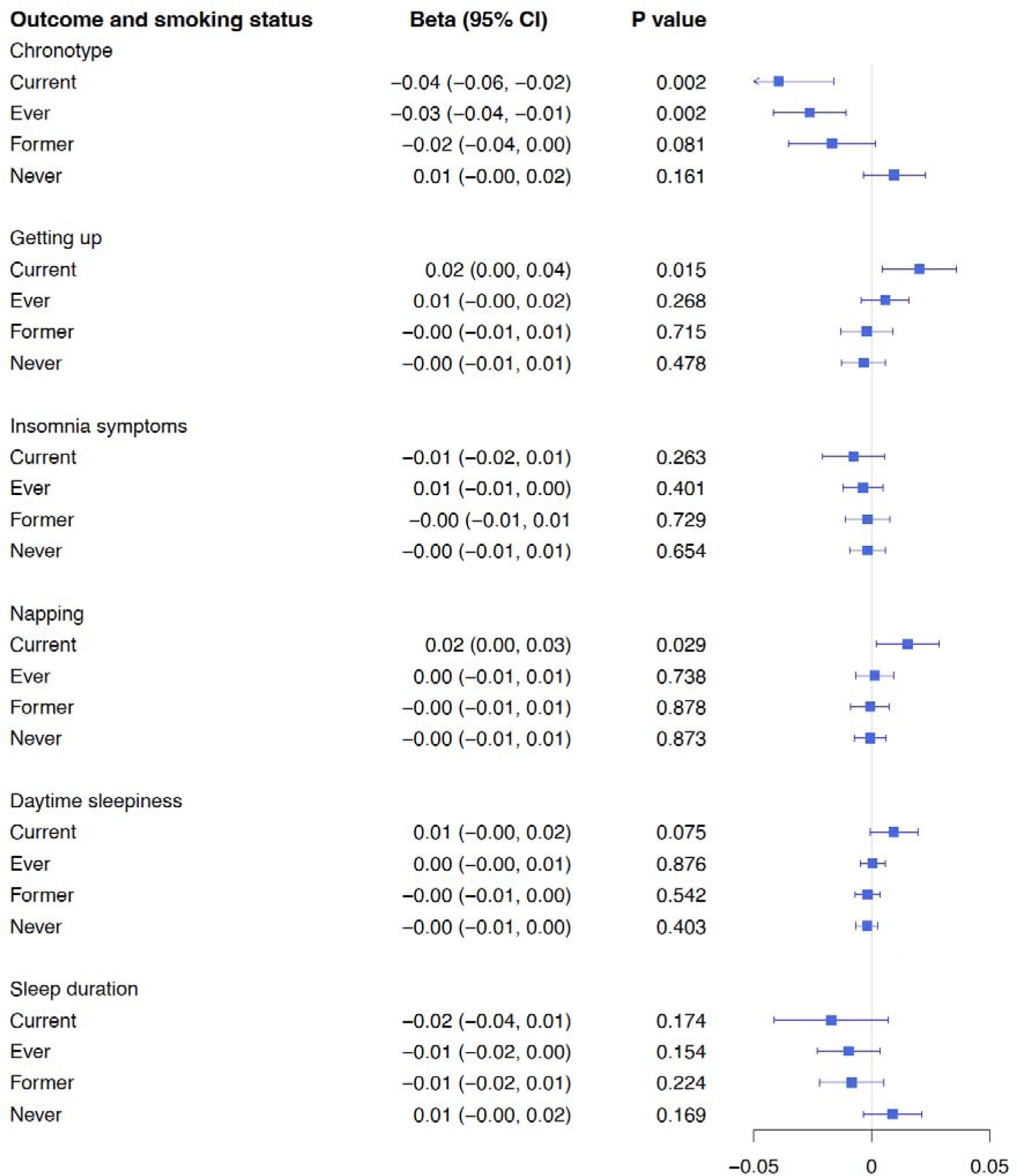
Forest plot of results from IVW-MVMR analysis of NMR on sleep outcomes measured in UK Biobank stratified by smoking status.

#### Direct effects of NMR on sleep outcomes when accounting for CPD

Using MVMR-IVW, we found evidence that increased NMR decreases the likelihood of being an evening person when accounting for CPD in current (β = −0.04, 95% confidence interval [95%CI] −0.06 to −0.02, p = 0.002) and ever smokers (β = −0.03, 95%CI −0.04 to −0.01, p = 0.003). We also observed evidence that increased NMR increases the likelihood of getting up in the morning in current smokers (β = 0.02, 95%CI 0.00 to 0.03, p = 0.015). We found evidence that increased NMR increased the likelihood of napping in current smokers (β = 0.02, 95%CI 0.00 to 0.03, p = 0.029). We also found suggestive evidence that increased NMR increased daytime sleepiness in current smokers (β = 0.01, 95%CI 0.00 to 0.02, p = 0.075).

We did not find evidence of consistent effects of NMR on insomnia symptoms or sleep duration in any of our MVMR analyses.

Effects observed for NMR were consistent in MVMR-Egger and we found limited evidence of association between genetically-proxied NMR and any of the sleep traits among never smokers, giving assurance against horizontal pleiotropy in the NMR genetic instrument (Supplementary Table S11).

### Direct effects of CPD on sleep outcomes when accounting for NMR

There was evidence that increased CPD increased the likelihood of being an evening person (β = 0.07, 95%CI 0.01 to 0.13, p = 0.035), increased the likelihood of napping (β = 0.04, 95%CI 0.00 to 0.07, p = 0.054 and decreased the likelihood of daytime sleepiness (β = −0.03, 95%CI −0.06 to 0.00, p = 0.052), when accounting for NMR in current smokers. There was some evidence that increased CPD increased sleep duration in former smokers (β = 0.05, 95%CI 0.01 to 0.09, p = 0.011). We did not find evidence of any effect of CPD on ease of getting up or insomnia symptoms, when accounting for NMR.

There was evidence of an inverse association between genetically-proxied CPD and evening chronotype in never smokers (β = −0.05, 95%CI −0.08 to −0.01, p = 0.008), indicative of horizontal pleiotropy (Supplementary Table S10). This association was not apparent in the MVMR-Egger analysis, while the positive effect of CPD on evening chronotype and inverse relationship between CPD and daytime sleepiness among current smokers were strengthened (Supplementary Table S11). The estimates for the effect of CPD on daytime napping among current smokers and of CPD on sleep duration among former smokers were consistent but with wider confidence intervals.

## Discussion

We used both univariable and multivariable Mendelian Randomization analyses to explore the total and direct effect of nicotine metabolite ratio (NMR) on sleep behaviours, accounting for cigarettes per day (CPD). When using outcome data stratified by smoking status in our MVMR analyses, we found evidence that increased NMR (i.e., lower levels of nicotine exposure per cigarette smoked) decreased the likelihood of being an evening person in current and ever smokers, increased the ease of getting up in the morning in current smokers and increased the likelihood of napping in current smokers, when accounting for smoking intensity. We did not find consistent effects of NMR on insomnia symptoms or sleep duration.

Previous studies have reported a higher prevalence of smoking among those with an evening chronotype (Ghotbi et al., 2023; Patterson et al., 2016). Further, a previous MR study showed that cigarettes per day increases the likelihood of being an evening person (Gibson et al., 2018). These support the findings from our study. Our novel finding is that a higher nicotine metabolite ratio (NMR) decreases the likelihood of being an evening person and increases the ease of getting up in the morning. This is important as it suggests, among smokers and for a given level of smoke exposure, those with a slower nicotine metabolism (i.e., higher circulating nicotine) might stay awake longer in the evening (due to nicotine arousal effects) while those with a higher nicotine metabolism (i.e. lower circulating nicotine) might wake up earlier (e.g., due to craving) (Ghotbi et al., 2023). We also found that a higher NMR was associated with an increased likelihood of napping and daytime sleepiness among current smokers, again suggesting less arousal in those with lower circulating nicotine.

While a previous study identified a causal link between smoking initiation and insomnia using Mendelian randomisation (Pasman et al., 2020), we did not find strong evidence for an effect of either smoking intensity or nicotine metabolism on insomnia symptoms among smokers.

Effects of both NMR and CPD on sleep traits were typically more prominent in current smokers than former smokers, suggesting that the effects of nicotine and smoking intensity on sleep traits are not long-lasting. In contrast to our findings, a recent study found that smoking cessation without nicotine replacement therapy did not affect chronotype, sleep quality or daytime sleepiness (Ghotbi et al., 2023). However, in that study sleep was assessed only 6 weeks after smoking cessation, which would not capture the long-term effects of cessation and the study was likely underpowered (n=49).

It is worth noting that our findings for the effects of CPD on some sleep outcomes (i.e. chronotype, napping, daytime sleepiness), once accounted for NMR, suggest there could be non-nicotine constituents that also impact sleep. However, further studies are required to isolate individual constituents and explore their direct effects on sleep outcomes while accounting for nicotine.

### Strengths and limitations

There are several strengths of this study, the first being the novel design of using NMR as a proxy measure for nicotine in multivariable MR analyses while accounting for CPD to isolate the effects of nicotine on sleep outcomes. Additionally, the large sample sizes of the GWAS for CPD and sleep outcomes used will have increased statistical power to detect effects. Further, the conditional F statistics indicated good instrument strength for both NMR and CPD in our MR and MVMR analyses. This is important for MR analyses, as weak instruments will give biased estimates (Sanderson et al., 2021).

There was some evidence for heterogeneity in the genetic instruments, especially for CPD, indicative of pleiotropy. However, because the effects were still observed in analyses using pleiotropy-robust methods (MR-Egger and MVMR-Egger), this suggests that pleiotropy is not driving these results. Further, effects estimates were consistently smaller for ever and former smokers, compared with current smokers, and we did not find any evidence of effects of NMR on any sleep outcomes in never smokers in MVMR analyses, providing additional reassurance against pleiotropy as this analysis serves as a negative control. While genetically-proxied CPD was related to chronotype among never smokers in both UV and MVMR, suggesting violation of the MR assumptions, this effect was not present in the MR-Egger analysis.

However, there are some important limitations to highlight. The small sample size for the NMR GWAS relative to CPD GWAS may have reduced power to detect direct effects of NMR on the sleep traits. However, conditional F statistics were sufficiently large and comparable between both NMR and CPD (Sanderson, Spiller & Bowden, 2021). Additionally, a more recent GWAS with a greater number of SNPs for CPD has since been released but this was not used as part of this study (Saunders et al., 2022). However, this may have led to even greater imbalance in the instrument strength for the two exposures, potentially leading to weak instrument bias.

Further, the self-report measures used to determine both smoking intensity and sleep behaviours may have been subject to measurement error (e.g., by participants misremembering or incorrectly reporting their behaviours)(Pierce & Burgess, 2013). Classical measurement error will typically result in bias towards the null in a two-sample MR framework, although this is not necessarily the case with multiple correlated exposures (Sanderson et al., 2019). MR performed on more refined smoking exposures (e.g. pack years) or sleep outcomes (e.g. derived from actigraphy devices or detailed sleep assessments) could yield more insights.

Another limitation is that it was outside of the scope of the current paper to evaluate the reciprocal effect of sleep behaviours on smoking intensity and nicotine metabolism, which has been previously investigated (Gibson et al., 2018; Pasman et al., 2020) and could have important implications, for example as smoking relapse is predicted by sleep disturbances following cessation (Patterson et al., 2019a).

It is also important to note that body mass index (BMI) has been shown to impact both sleep (Garfield, 2019) and smoking behaviours (Carreras-Torres et al., 2018), but it was not specifically examined in this study. Although BMI was controlled for in the NMR GWAS, it was not controlled for in the CPD or sleep GWAS. While a trivariate MR analysis to account for the role of BMI would be beneficial, this would further reduce the conditional instrument strength in the MVMR model and so was not attempted here.

Finally, the data used as part of this analysis were all based on cohorts with European ancestry, meaning these results cannot be generalised to populations with non-European ancestry.

### Conclusion

In conclusion, we found evidence that increased NMR (which equates to decreased nicotine exposure per fixed level of smoke exposure) decreases the likelihood of being an evening person and increases ease of getting up in the morning. We also found evidence that increased NMR increases the likelihood of napping and to a lesser extent, daytime sleepiness. These findings suggest that nicotine plays a role in the relationship between smoking and certain sleep behaviours. They also suggest that nicotine could impact certain traits independently from the remaining constituents of tobacco smoke, which has implications for individuals using nicotine replacement therapy and nicotine delivery systems such as e-cigarettes. However, the limited evidence for an impact of NMR on insomnia suggests that nicotine itself does not strongly impact more problematic sleep behaviours. Overall, our study contributes to evidence regarding the effects of smoking on circadian sleep-wake behaviour. Future studies would benefit from using larger datasets on NMR and more refined measures of smoking and sleep to help strengthen our inference in these relationships.

## Data Availability

All data produced in the present work are contained in the manuscript.

