## Supplementary Figures for "Exploring the role of nicotine and smoking in sleep behaviours: A multivariable Mendelian Randomisation study"

Supplementary Figure S1. Single SNP analysis from UVMR of NMR on chronotype UK Biobank stratified by smoking status

a. Current smokers

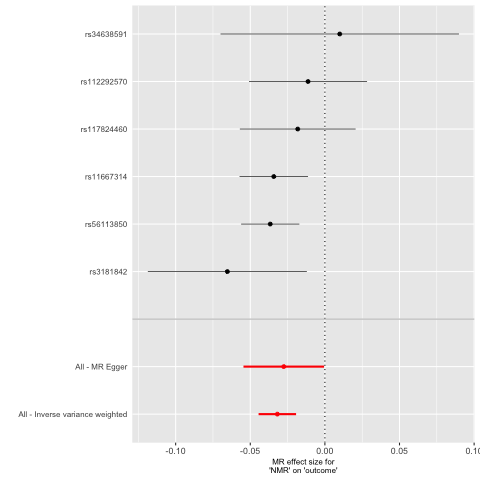

b. Ever smokers

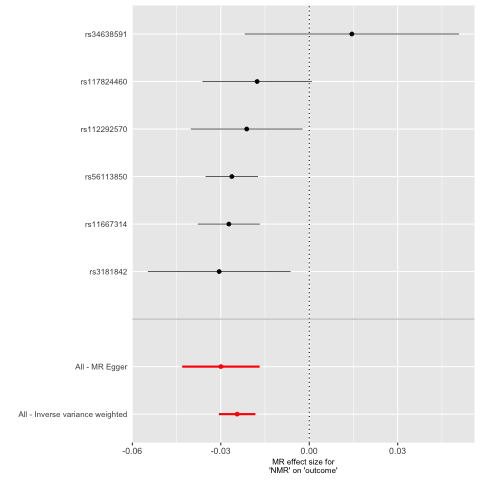

c. Former smokers

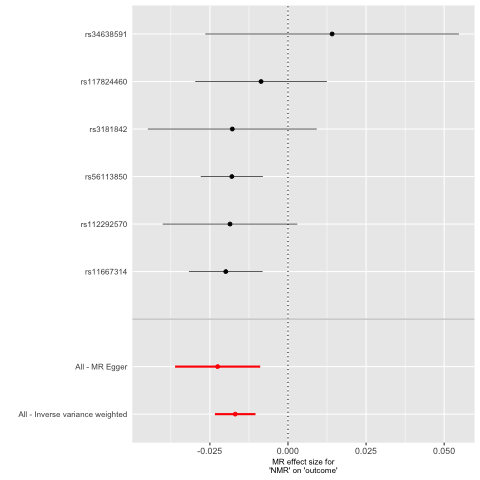

d. Never smokers

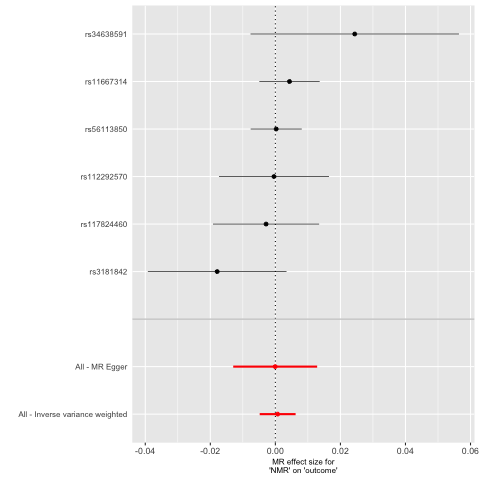

Supplementary Figure 2. Funnel plot from UVMR of NMR on chronotype UK Biobank stratified by smoking status

a. Current smokers

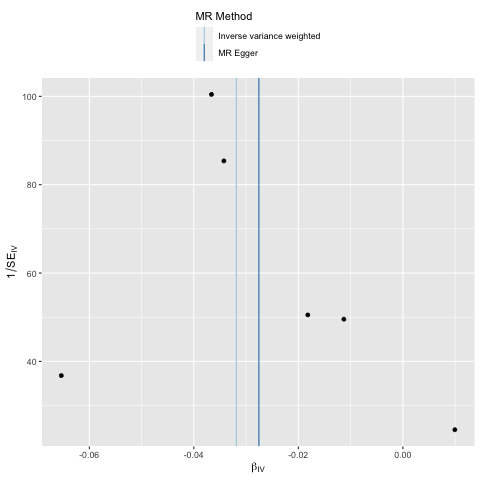

b. Ever smokers

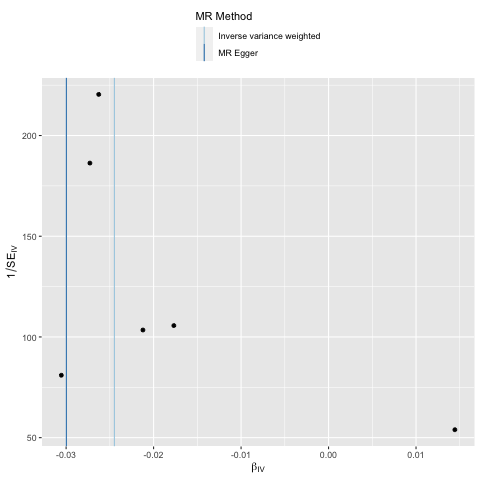

c. Former smokers

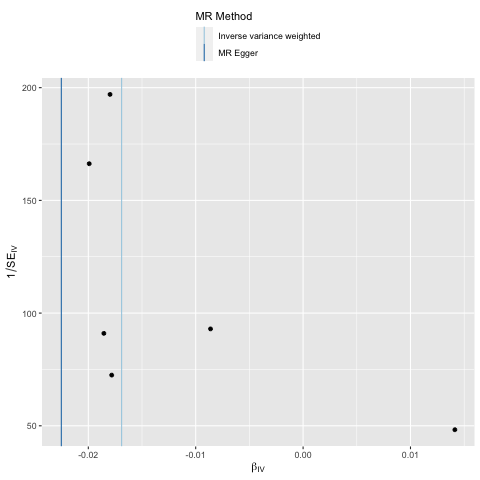

d. Never smokers

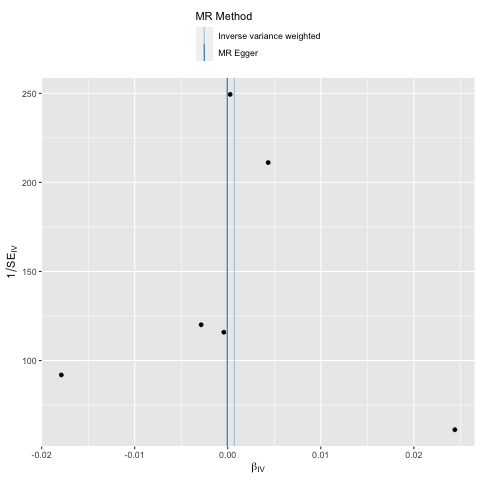

Supplementary Figure 3. Leave one out analysis from UVMR of NMR on chronotype UK Biobank stratified by smoking status

a. Current smokers

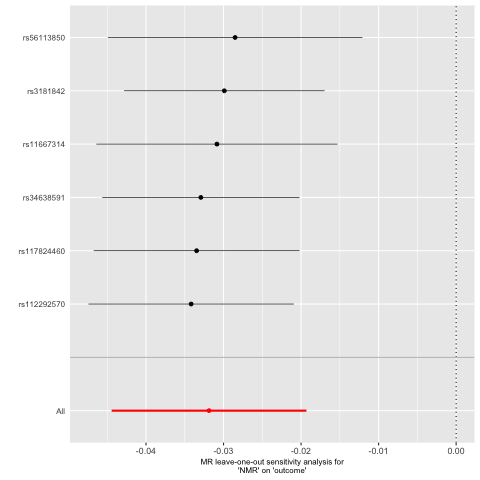

b. Ever smokers

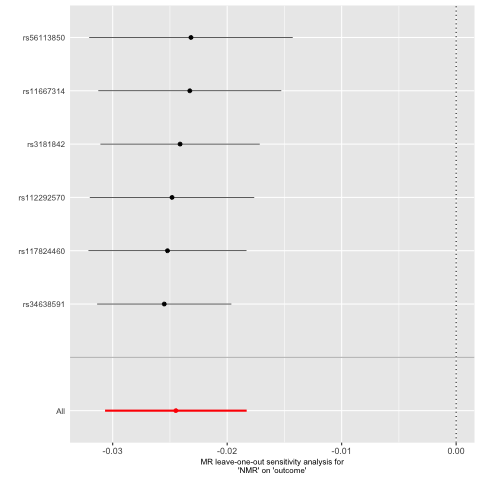

c. Former smokers

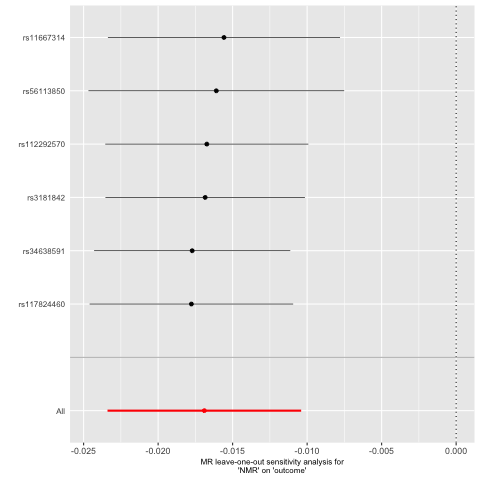

d. Never smokers

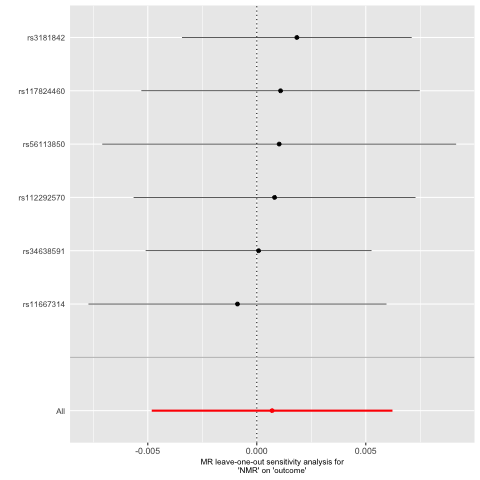

Supplementary Figure 4. Single SNP analysis from UVMR of NMR on getting up UK Biobank stratified by smoking status

a. Current smokers

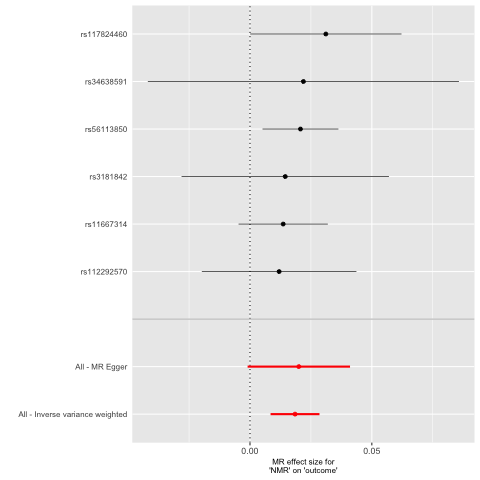

b. Ever smokers

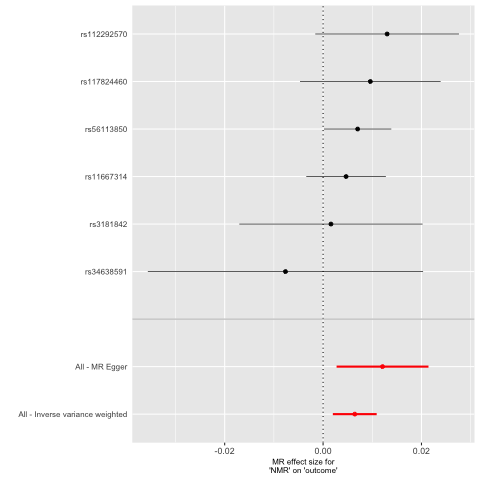

c. Former smokers

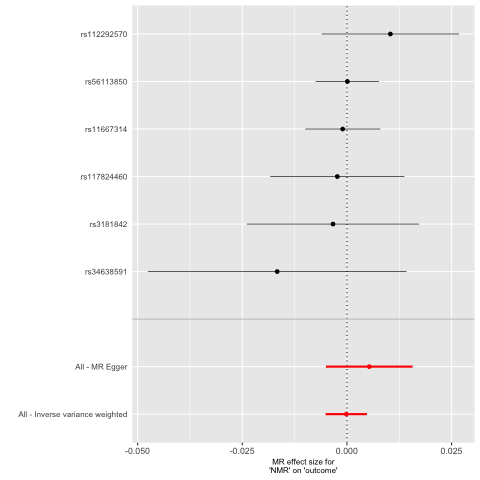

d. Never smokers

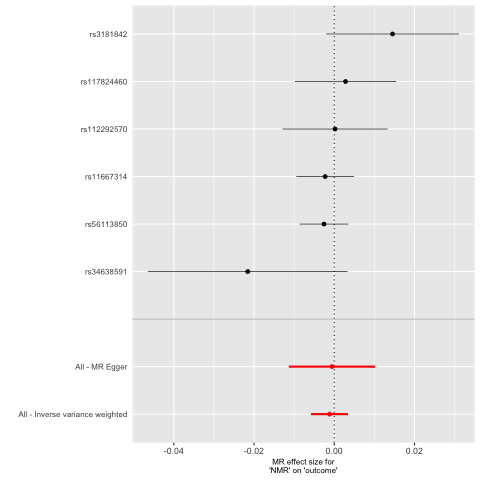

Supplementary Figure 5. Funnel plot from UVMR of NMR on getting up UK Biobank stratified by smoking status

a. Current smokers

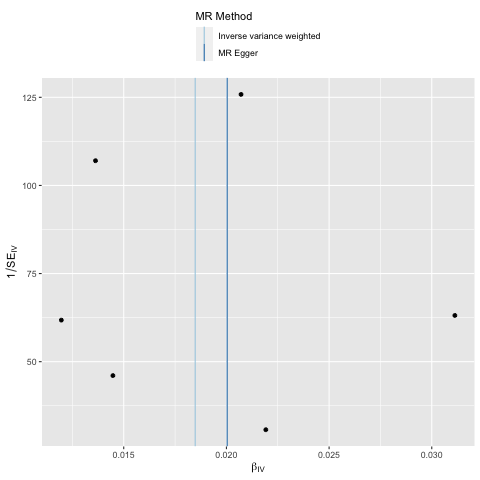

b. Ever smokers

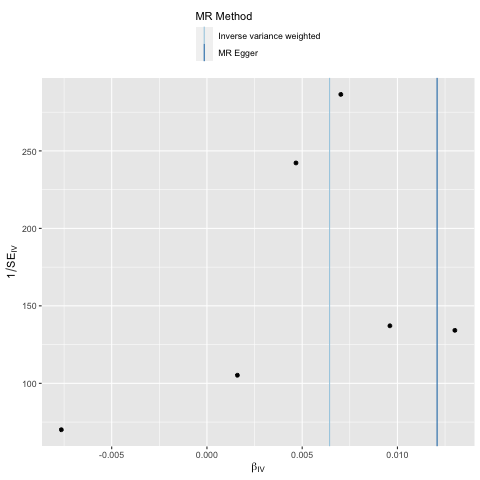

c. Former smokers

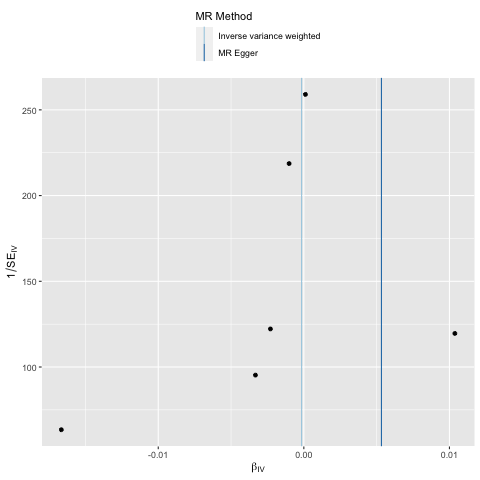

d. Never smokers

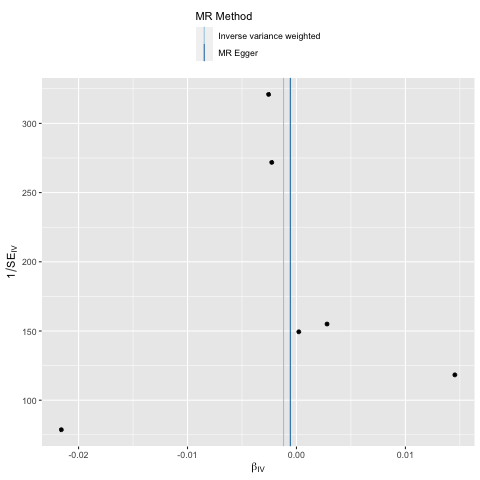

Supplementary Figure 6. Leave one out analysis from UVMR of NMR on getting up UK Biobank stratified by smoking status

a. Current smokers

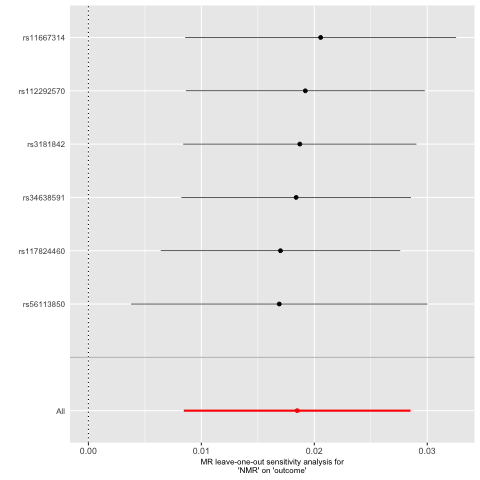

b. Ever smokers

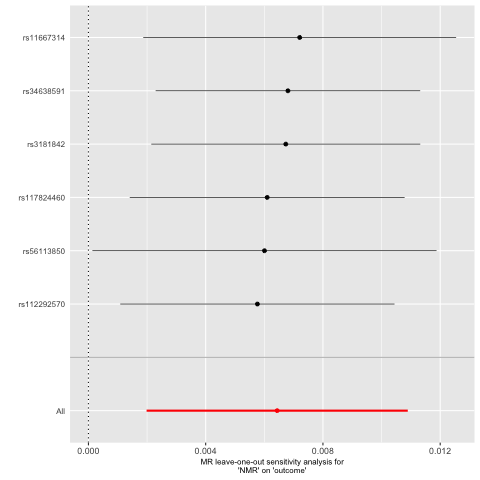

c. Former smokers

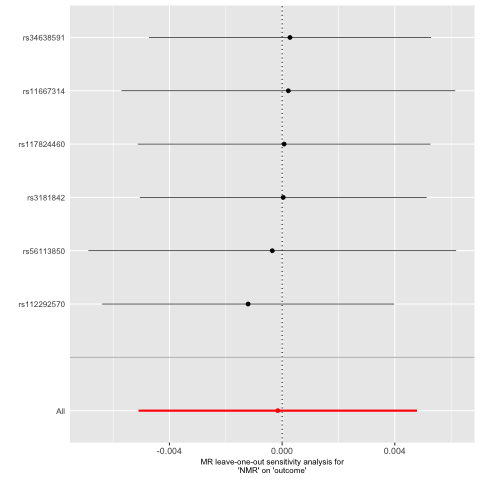

d. Never smokers

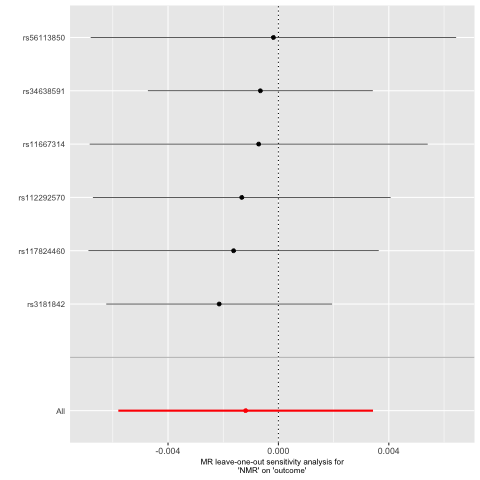

Supplementary Figure 7. Single SNP analysis from UVMR of NMR on insomnia symptoms UK Biobank stratified by smoking status

a. Current smokers

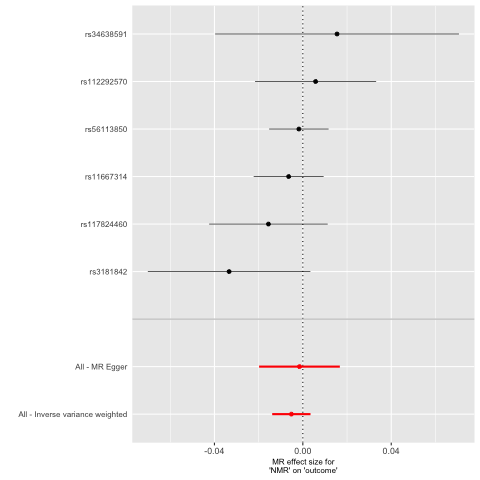

b. Ever smokers

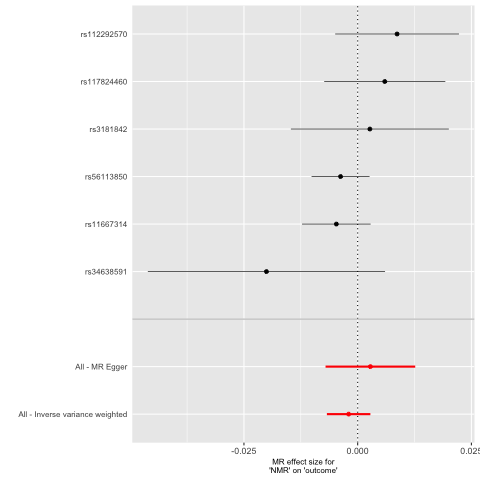

c. Former smokers

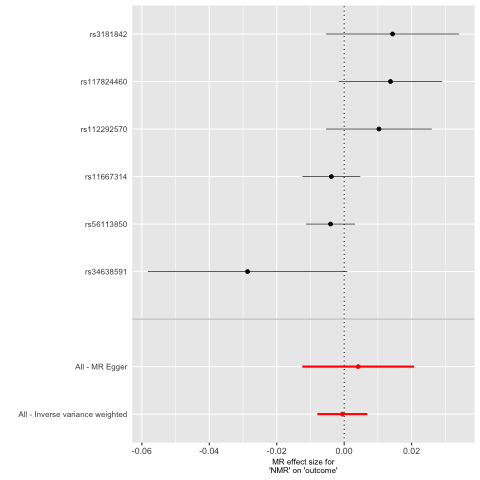

d. Never smokers

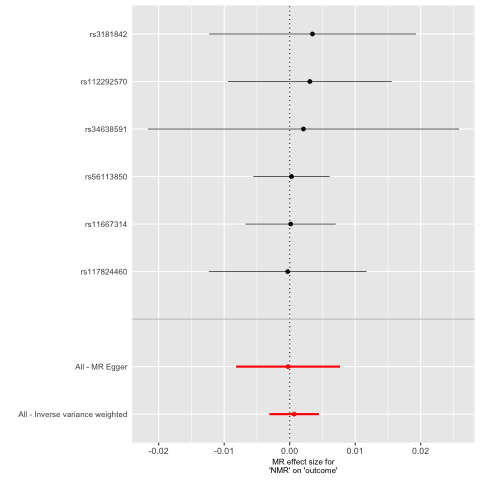

Supplementary Figure 8. Funnel plot from UVMR of NMR on insomnia symptoms UK Biobank stratified by smoking status

a. Current smokers

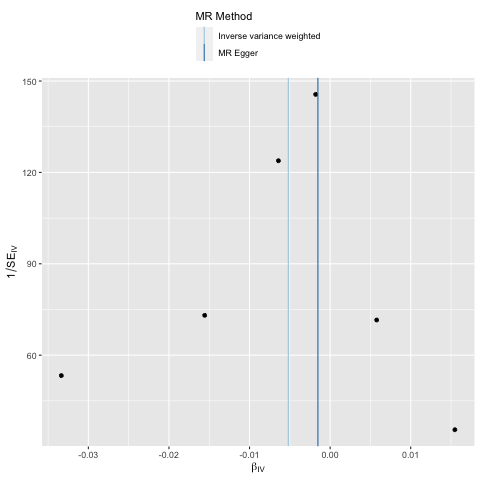

b. Ever smokers

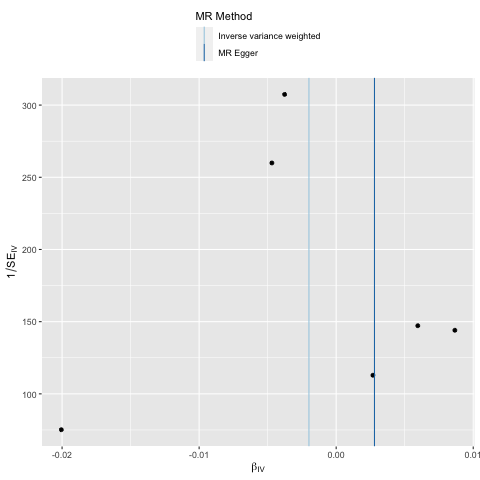

c. Former smokers

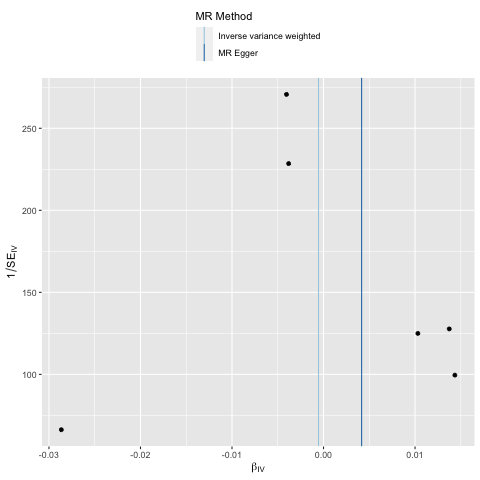

d. Never smokers

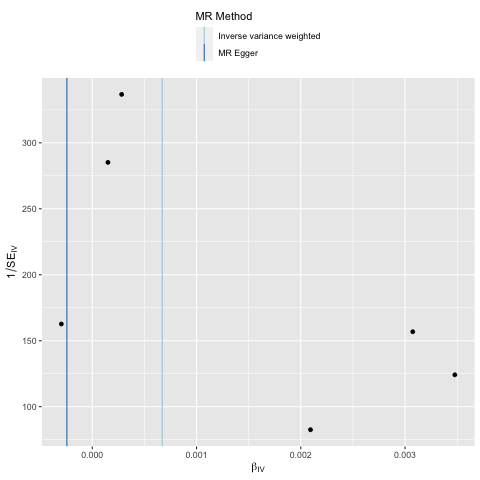

Supplementary Figure 9. Leave one out analysis from UVMR of NMR on insomnia symptoms UK Biobank stratified by smoking status

a. Current smokers

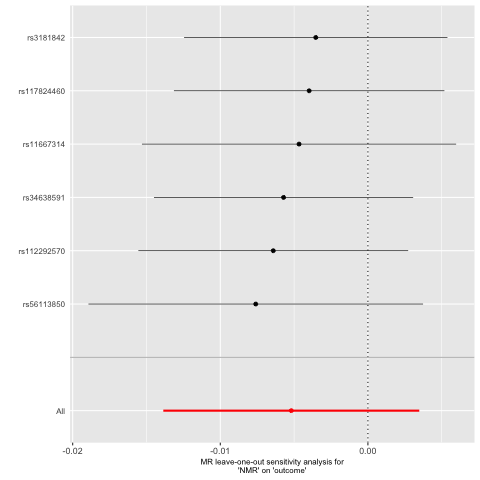

b. Ever smokers

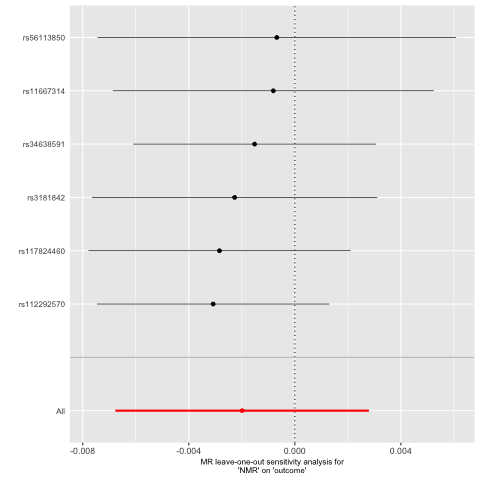

c. Former smokers

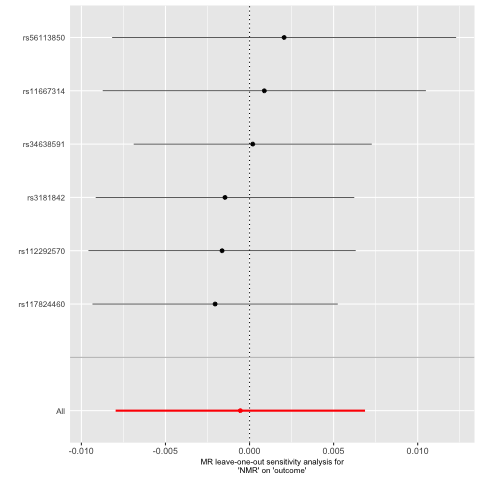

d. Never smokers

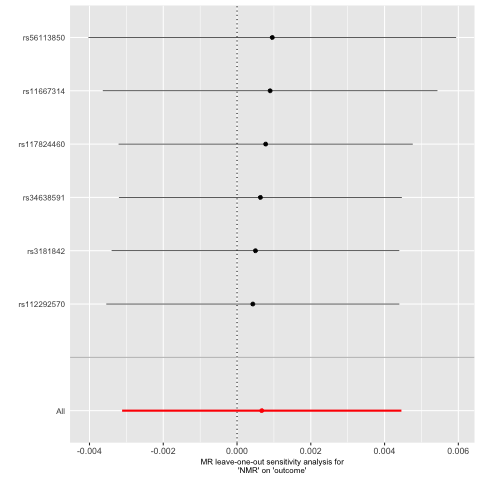

Supplementary Figure 10. Single SNP analysis from UVMR of NMR on napping UK Biobank stratified by smoking status

a. Current smokers

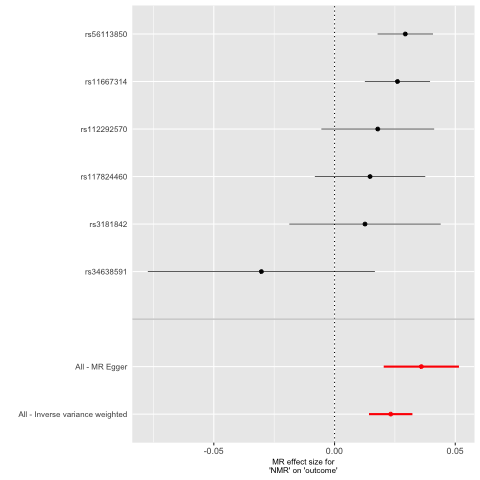

b. Ever smokers

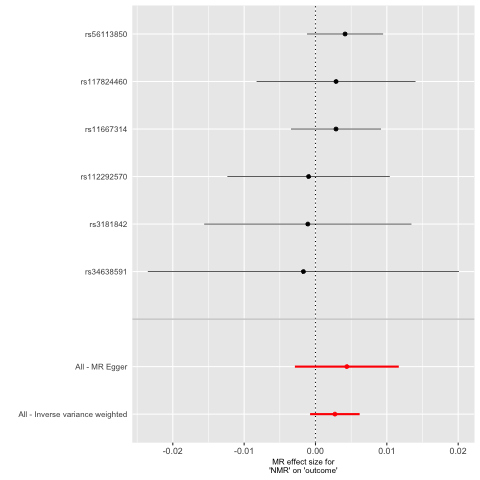

c. Former smokers

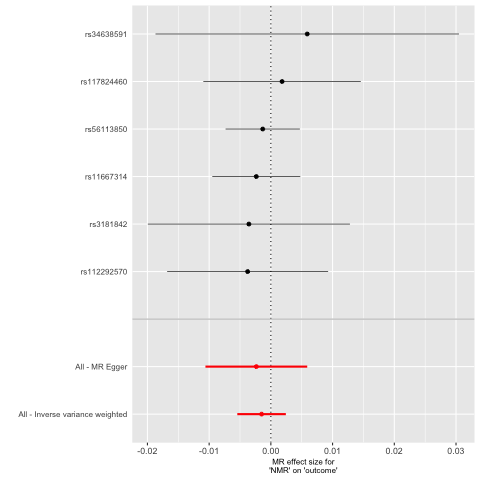

d. Never smokers

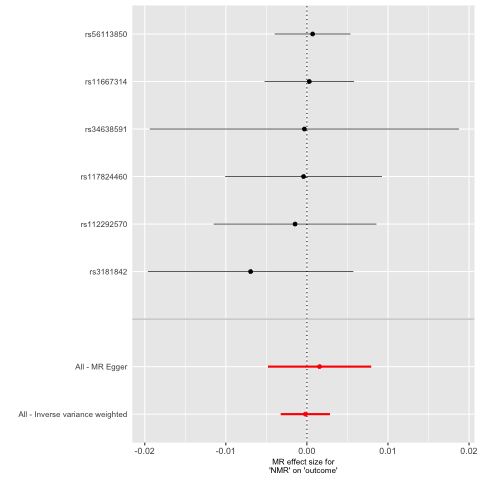

Supplementary Figure 11. Funnel plot from UVMR of NMR on napping UK Biobank stratified by smoking status

a. Current smokers

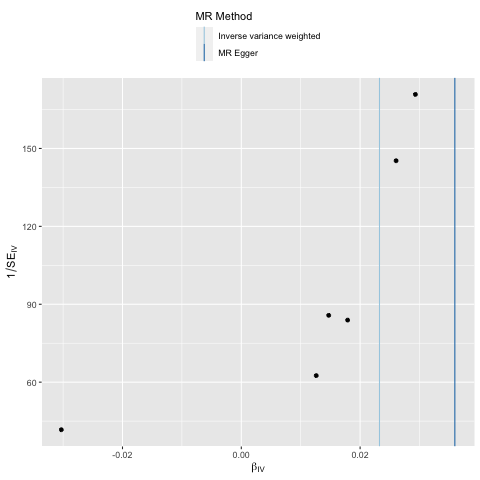

b. Ever smokers

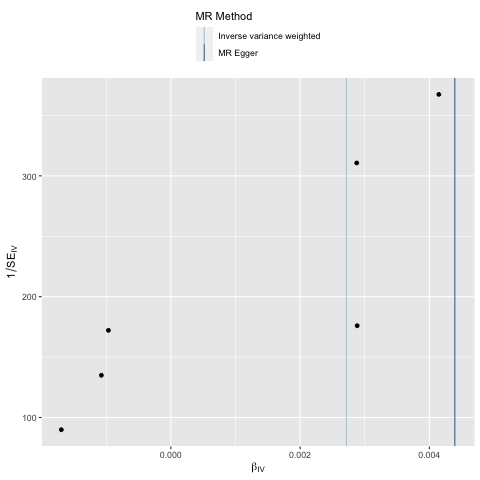

c. Former smokers

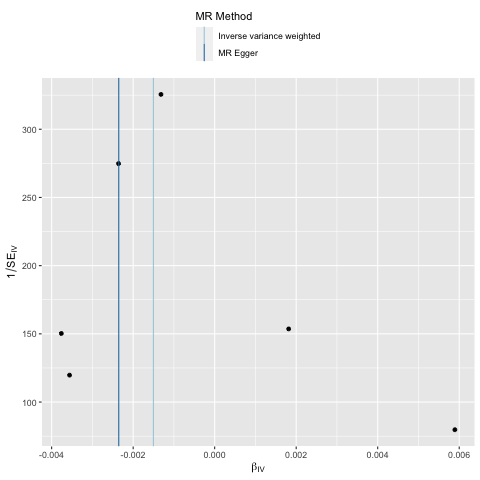

d. Never smokers

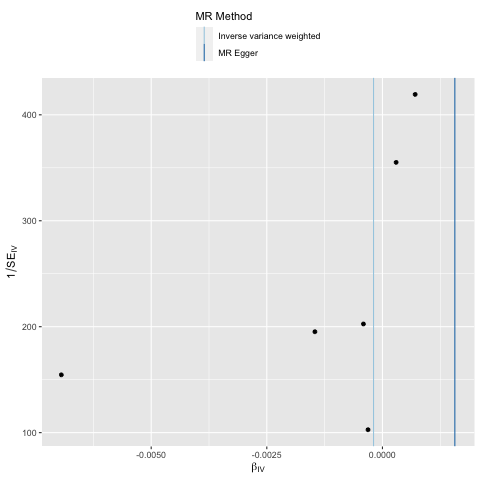

Supplementary Figure 12. Leave one out analysis from UVMR of NMR on napping UK Biobank stratified by smoking status

a. Current smokers

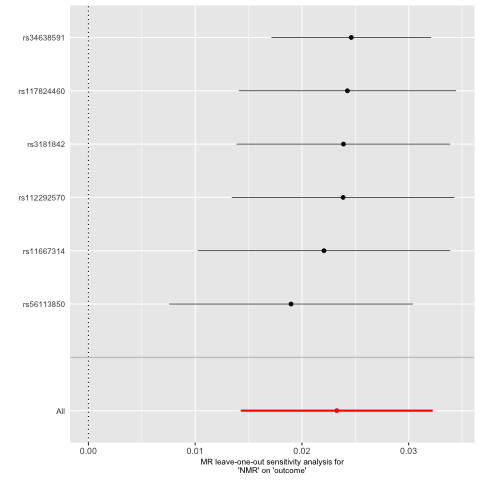

b. Ever smokers

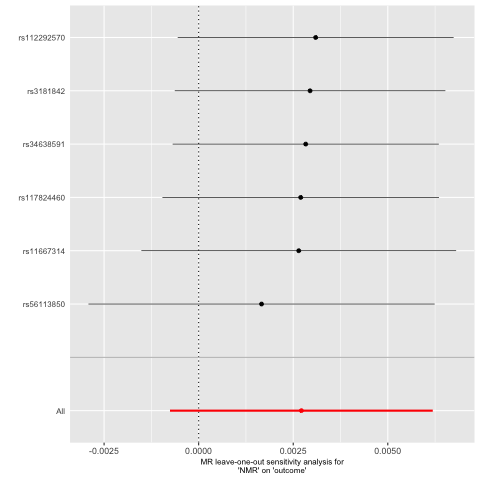

c. Former smokers

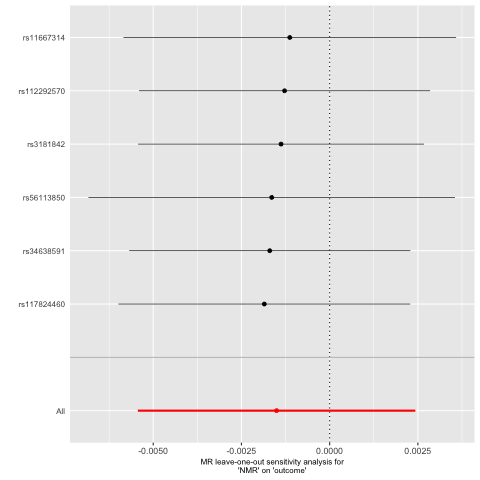

d. Never smokers

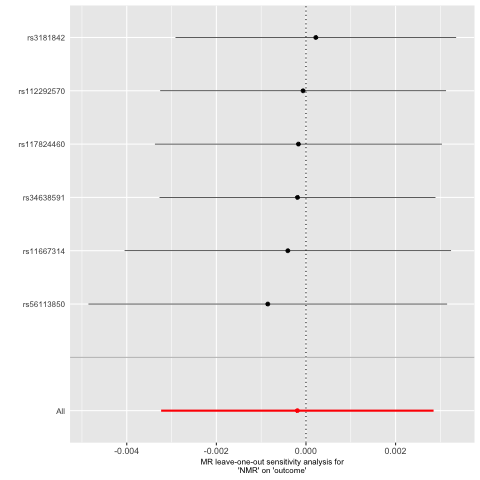

Supplementary Figure 13. Single SNP analysis from UVMR of NMR on daytime sleepiness UK Biobank stratified by smoking status

a. Current smokers

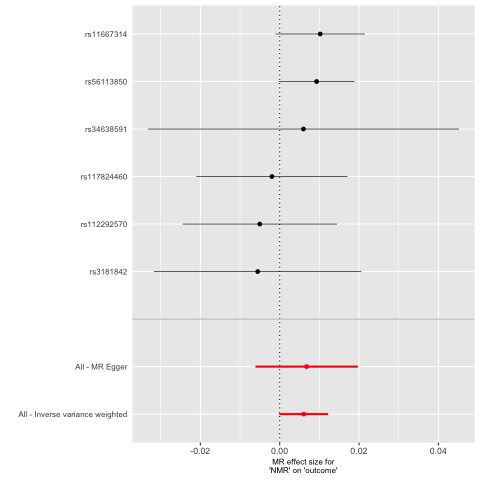

b. Ever smokers

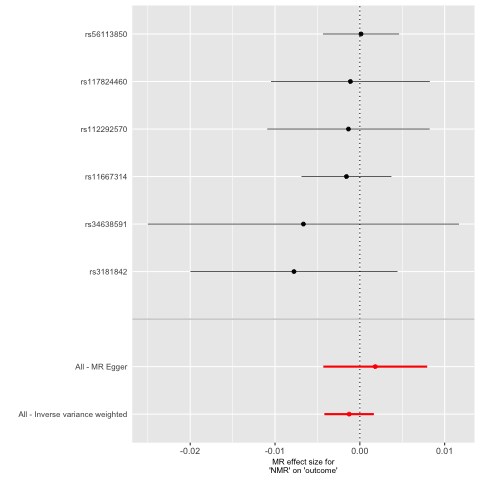

c. Former smokers

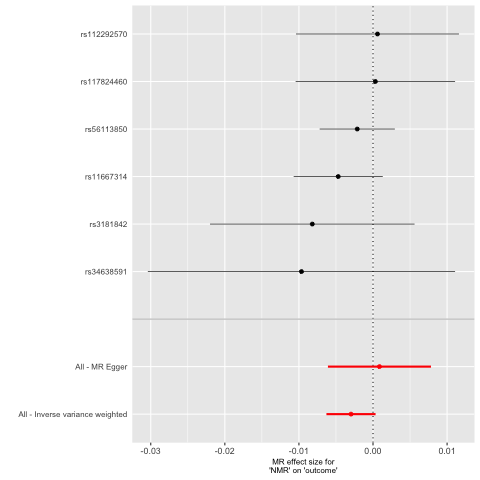

d. Never smokers

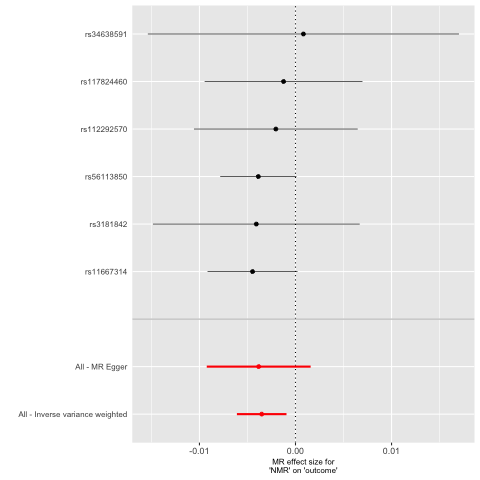

Supplementary Figure 14. Funnel plot from UVMR of NMR on daytime sleepiness UK Biobank stratified by smoking status

a. Current smokers

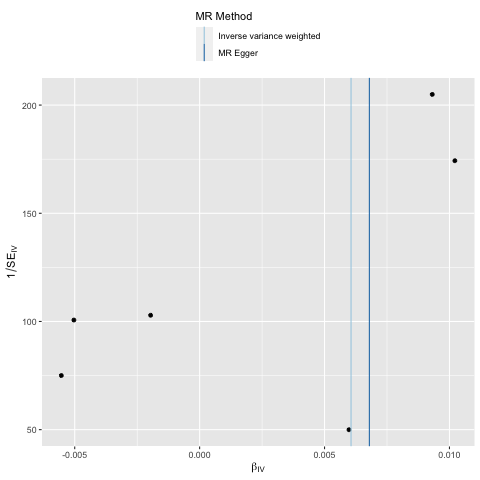

b. Ever smokers

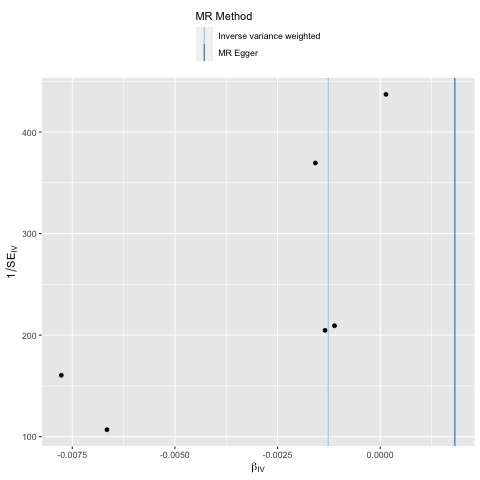

c. Former smokers

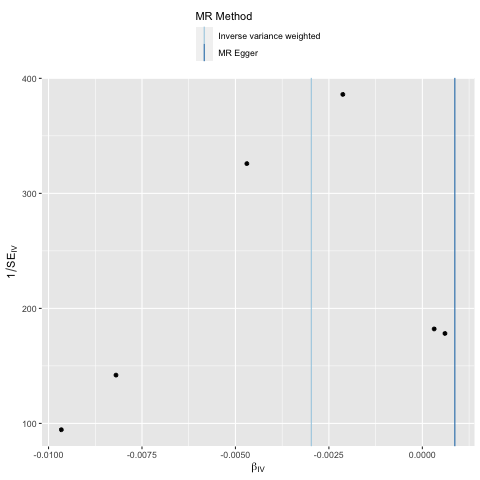

d. Never smokers

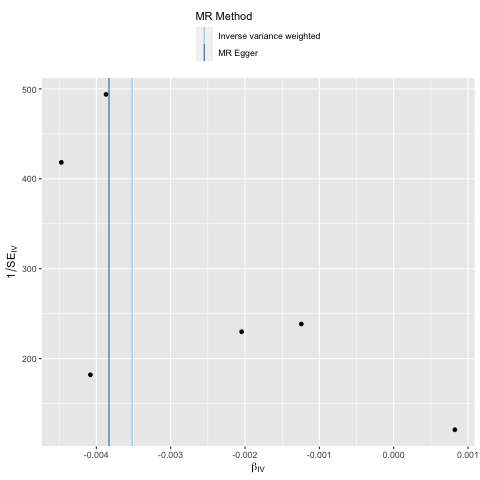

Supplementary Figure 15. Leave one out analysis from UVMR of NMR on daytime sleepiness UK Biobank stratified by smoking status

a. Current smokers

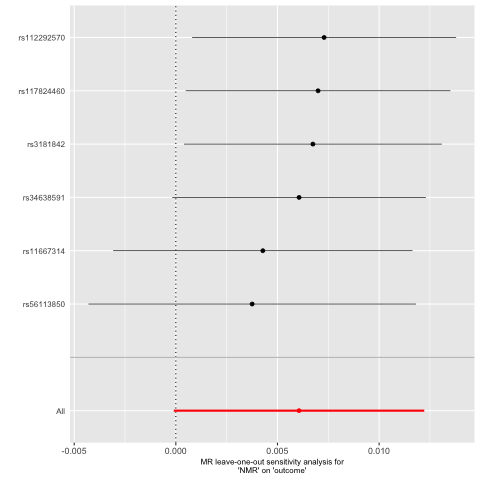

b. Ever smokers

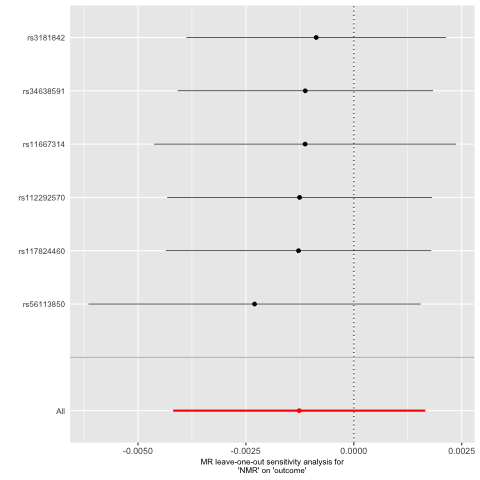

c. Former smokers

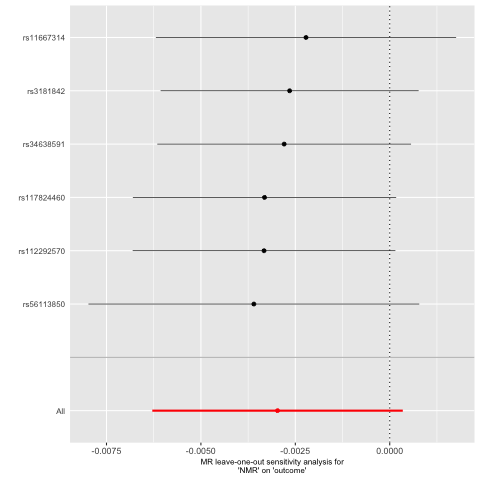

d. Never smokers

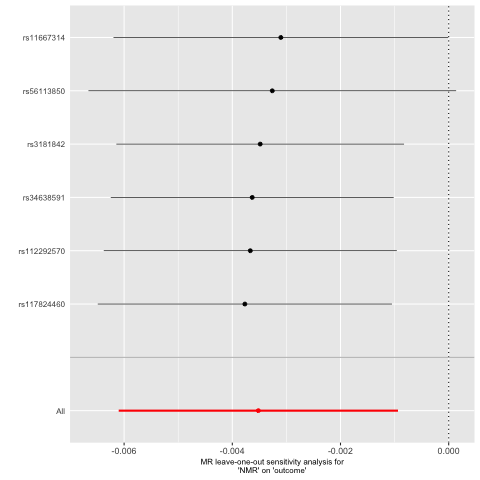

Supplementary Figure 16. Single SNP analysis from UVMR of NMR on sleep duration UK Biobank stratified by smoking status

a. Current smokers

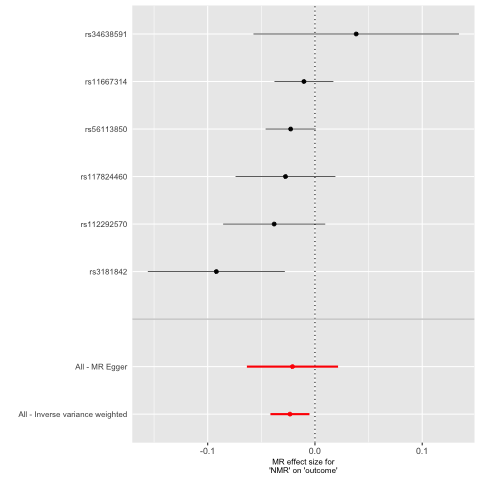

b. Ever smokers

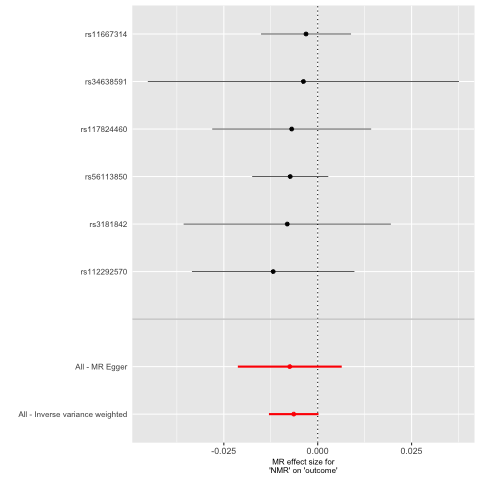

c. Former smokers

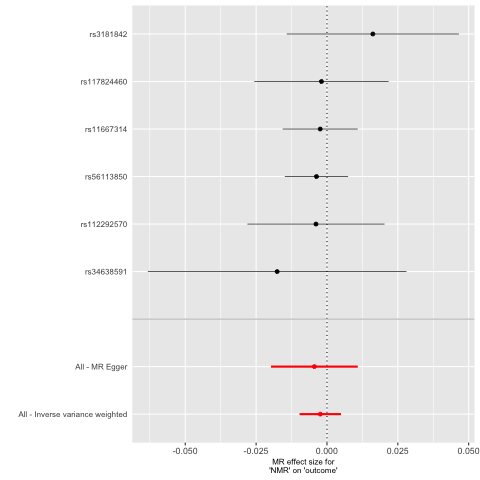

d. Never smokers

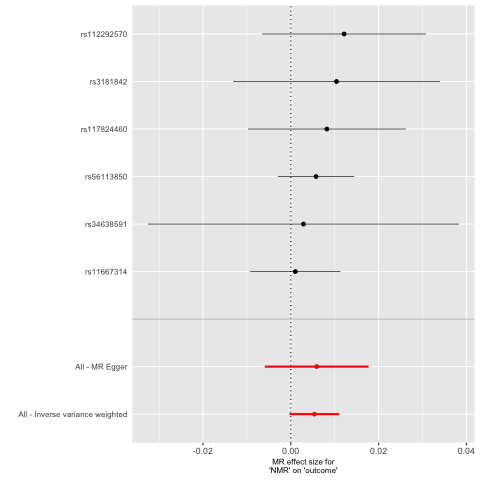

Supplementary Figure 17. Funnel plot from UVMR of NMR on sleep duration UK Biobank stratified by smoking status

a. Current smokers

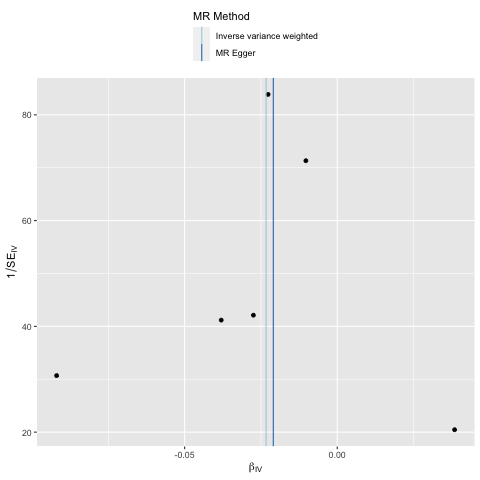

b. Ever smokers

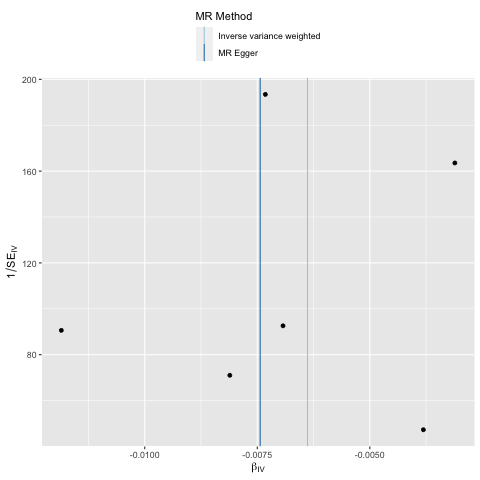

c. Former smokers

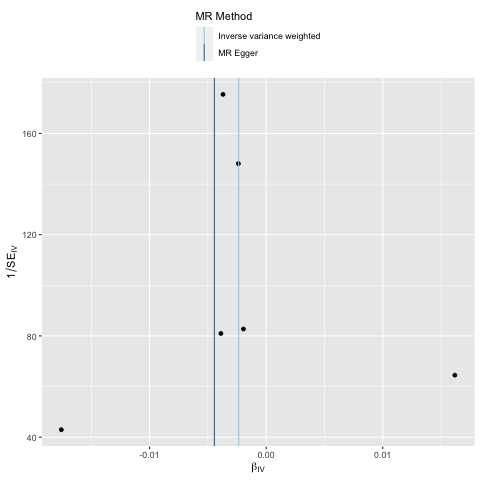

d. Never smokers

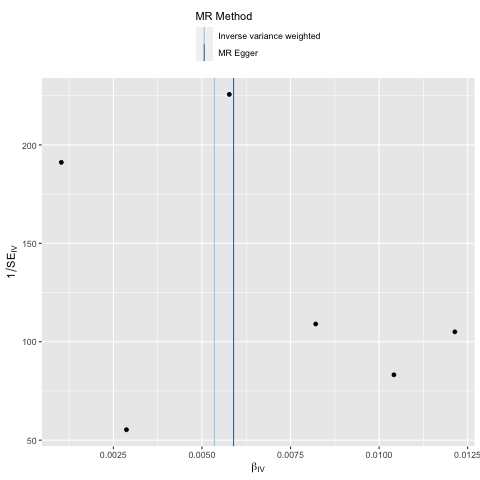

Supplementary Figure 18. Leave one out analysis from UVMR of NMR on sleep duration UK Biobank stratified by smoking status

a. Current smokers

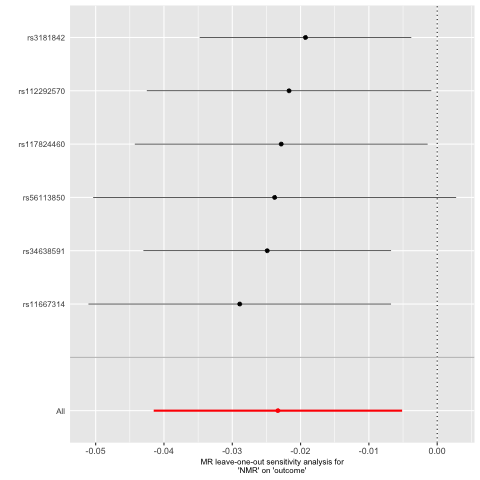

b. Ever smokers

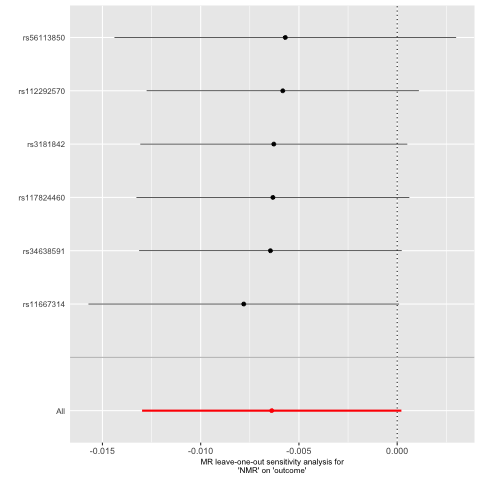

c. Former smokers

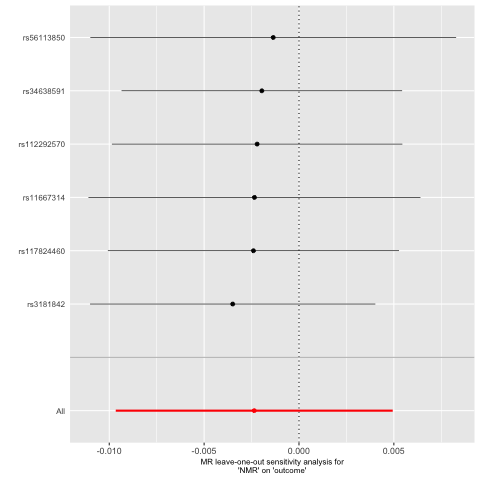

d. Never smokers

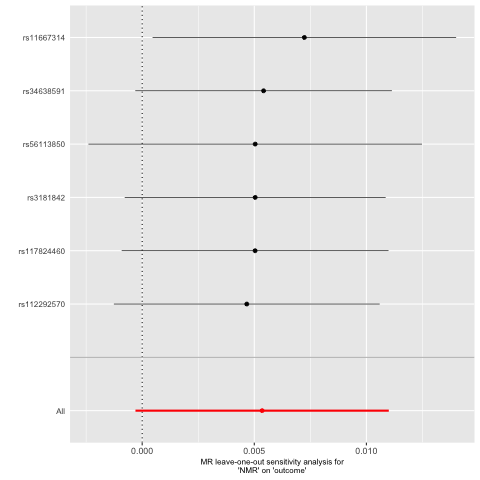

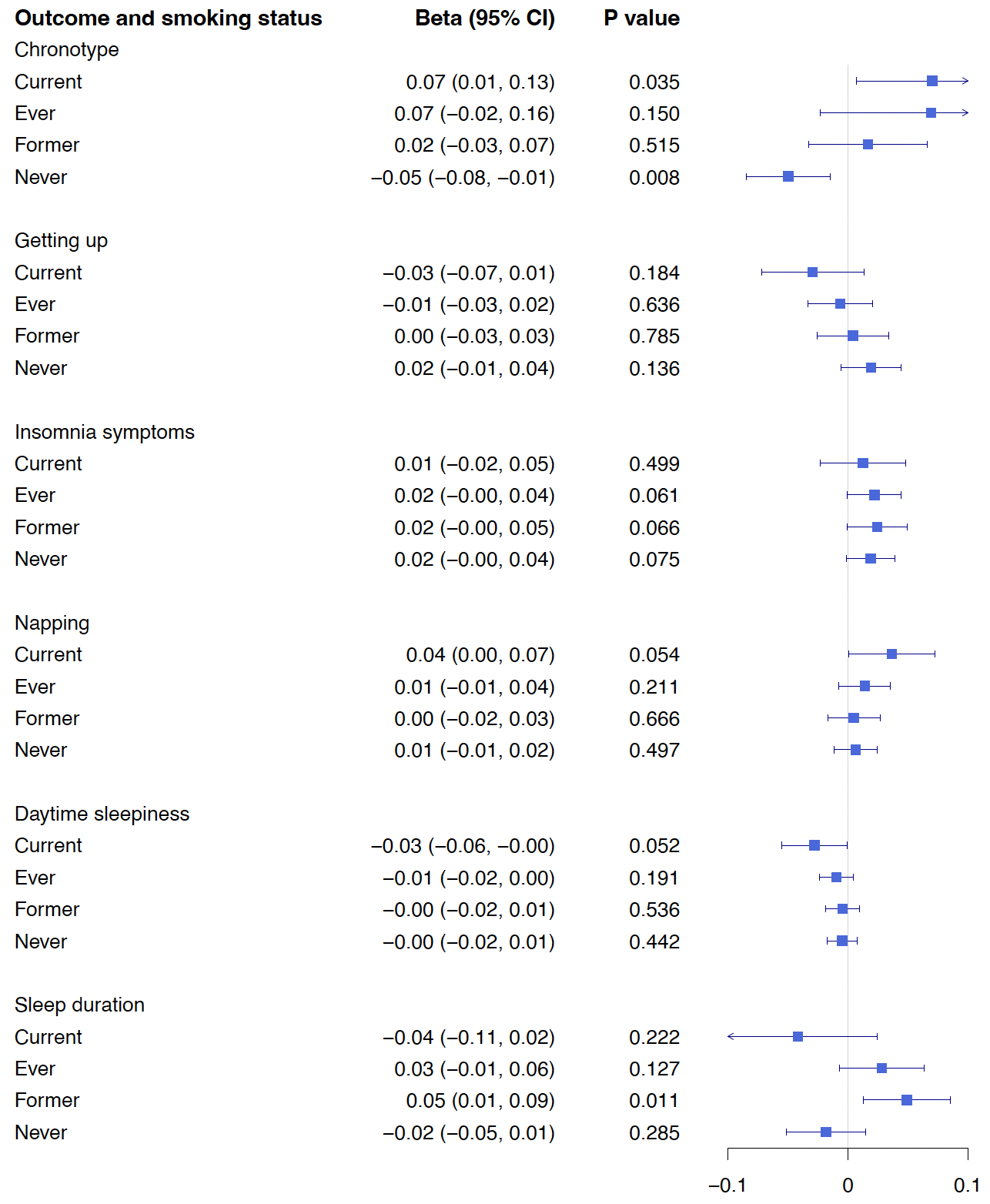

Supplementary Figure 19. Forest plot of results from IVW-MVMR analysis of CPD and sleep outcomes measured in UK Biobank stratified by smoking status
