## Supplementary Tables for "Exploring the role of nicotine and smoking in sleep behaviours: A multivariable Mendelian Randomisation study"

| **Outcome** | **Mean** | **Standard Deviation (SD)** |
| --- | --- | --- |
| Smoking status |  |  |
| **Chronotype** |  |  |
| Current | 2.44 | 1.00 |
| Ever | 2.26 | 0.96 |
| Former | 2.21 | 0.93 |
| Never | 2.14 | 0.91 |
| **Getting up** |  |  |
| Current | 2.98 | 0.86 |
| Ever | 3.11 | 0.79 |
| Former | 3.14 | 0.77 |
| Never | 3.12 | 0.76 |
| **Insomnia** |  |  |
| Current | 2.07 | 0.73 |
| Ever | 2.08 | 0.72 |
| Former | 2.08 | 0.72 |
| Never | 2.02 | 0.72 |
| **Napping** |  |  |
| Current | 1.57 | 0.63 |
| Ever | 1.53 | 0.61 |
| Former | 1.52 | 0.61 |
| Never | 1.45 | 0.58 |
| **Narcolepsy** |  |  |
| Current | 0.27 | 0.52 |
| Ever | 0.27 | 0.51 |
| Former | 0.27 | 0.50 |
| Never | 0.25 | 0.49 |
| **Sleep duration** |  |  |
| Current | 7.09 | 1.26 |
| Ever | 7.16 | 1.14 |
| Former | 7.18 | 1.10 |
| Never | 7.17 | 1.05 |

Supplementary Table S1. Mean and standard deviations for outcome responses

Supplementary Table S2. Response codes for sleep behaviours measured in UK Biobank

| **Sleep behaviour** | **N** | **Question** | **Response** | **Code** |
| --- | --- | --- | --- | --- |
| Morning/evening person (chronotype) | 413,343 | Do you consider yourself to be (choose from options): | 1 | Definitely a 'morning' person |
|  |  |  | 2 | More 'morning' than 'evening' person |
|  |  |  | 3 | More 'evening' than 'morning' person |
|  |  |  | 4 | Definitely an evening person |
| Getting up in the morning | 461,658 | On an average day, how easy do you find getting up in the morning? | 1 | Not at all easy |
|  |  |  | 2 | Not very easy |
|  |  |  | 3 | Fairly easy |
|  |  |  | 4 | Very easy |
| Insomnia symptoms | 462,341 | Do you have trouble falling asleep at night or do you wake up in the middle of the night? | 1 | Never/rarely |
|  |  |  | 2 | Sometimes |
|  |  |  | 3 | Usually |
| Napping during the day | 462,400 | Do you have a nap during the day? | 1 | Never/rarely |
|  |  |  | 2 | Sometimes |
|  |  |  | 3 | Usually |
| Daytime sleepiness | 460,913 | How likely are you to doze off or fall asleep during the daytime when you don't mean to? (e.g. when working, reading or driving) | 1 | Never/rarely |
|  |  |  | 2 | Sometimes |
|  |  |  | 3 | Often |
|  |  |  | 4 | All of the time |
| Sleep duration | 460,099 | About how many hours sleep do you get in every 24 hours? (please include naps) | *** | N/A |
| *** Participants were asked to self-report the number of hours rather than choose from options | | |  |  |

Supplementary Table S3. Univariable Mendelian randomisation analysis (IVW-MR) of nicotine metabolite ratio (NMR), cigarettes per day (CPD) and continuous sleep outcomes measured in UK Biobank stratified by smoking status

| Sleep outcome | Smoking status |  | NMR |  |  | CPD |  |
| --- | --- | --- | --- | --- | --- | --- | --- |
|  |  | Effect | 95% CI | P-value | Effect | 95% CI | P-value |
| Chronotype* | Current | -0.03 | -0.04, -0.02 | 0.006 | 0.13 | 0.09, 0.20 | 4.94x10^-10^ |
|  | Ever | -0.02 | -0.03, -0.02 | 8.59x10^-15^ | 0.07 | 0.04, 0.10 | 2.79x10^-06^ |
|  | Former | -0.02 | -0.02, -0.01 | 3.45x10^-07^ | 0.04 | 0.02, 0.07 | 0.003 |
|  | Never | 0.00 | -0.00, 0.01 | 0.803 | -0.04 | -0.06, -0.02 | 5.12x10^-05^ |
| Getting up | Current | 0.02 | 0.01, 0.03 | <0.001 | -0.05 | -0.07, -0.02 | 0.001 |
|  | Ever | 0.01 | 0.00, 0.01 | 0.005 | -0.02 | -0.04, -0.01 | 0.005 |
|  | Former | -0.00 | -0.01, 0.00 | 0.951 | -0.01 | -0.03, 0.00 | 0.107 |
|  | Never | -0.00 | -0.01, 0.00 | 0.614 | 0.02 | 0.01, 0.04 | 0.002 |
| Insomnia symptoms | Current | -0.01 | -0.01, 0.00 | 0.241 | 0.02 | -0.00, 0.04 | 0.107 |
|  | Ever | -0.00 | -0.01, 0.00 | 0.415 | 0.01 | -0.00, 0.02 | 0.134 |
|  | Former | -0.00 | -0.01, 0.01 | 0.885 | 0.01 | -0.01, 0.02 | 0.289 |
|  | Never | 0.00 | -0.00, 0.00 | 0.728 | 0.00 | -0.01, 0.01 | 0.867 |
| Napping | Current | 0.02 | 0.01, 0.03 | 3.95x10^-07^ | 0.02 | 0.00, 0.04 | 0.023 |
|  | Ever | 0.00 | -0.00, 0.01 | 0.125 | 0.01 | -0.00, 0.02 | 0.104 |
|  | Former | -0.00 | -0.01, 0.00 | 0.434 | 0.01 | -0.01, 0.02 | 0.378 |
|  | Never | -0.00 | -0.00, 0.00 | 0.901 | 0.01 | -0.00, 0.02 | 0.148 |
| Daytime sleepiness | Current | 0.01 | -10.00, 0.01 | 0.054 | -0.05 | -0.07, -0.03 | 1.61x10^-06^ |
|  | Ever | -0.00 | -0.00, 0.00 | 0.396 | -0.02 | -0.02, -0.01 | <0.001 |
|  | Former | -0.00 | -0.01, 0.00 | 0.079 | -0.01 | -0.02, 0.00 | 0.090 |
|  | Never | -0.00 | -0.01, -0.00 | 0.01 | -0.00 | -0.01, 0.00 | 0.276 |
| Sleep duration | Current | -0.02 | -0.04, -0.01 | 0.012 | -0.01 | -0.05, 0.02 | 0.500 |
|  | Ever | -0.00 | -0.01, 0.00 | 0.058 | 0.04 | 0.09, 0.60 | <0.001 |
|  | Former | -0.00 | -0.01, 0.00 | 0.527 | 0.06 | 0.03, 0.08 | 1.45x10^-07^ |
|  | Never | 0.01 | -0.00, 0.01 | 0.063 | -0.02 | -0.04, 0.00 | 0.103 |

*Coded towards being an evening person

Supplementary Table S4. Univariable Mendelian randomisation analysis (MR-Egger) of nicotine metabolite ratio (NMR), cigarettes per day (CPD) and continuous sleep outcomes measured in UK Biobank stratified by smoking status

| Sleep outcome | Smoking status | NMR | | | CPD | | |
| --- | --- | --- | --- | --- | --- | --- | --- |
|  |  | Effect | 95% CI | P-value | Effect | 95% CI | P-value |
| Chronotype | Current | -0.03 | -0.05, -0.00 | 0.116 | 0.19 | 0.13, 0.24 | 5.54x10^-08^ |
|  | Ever | -0.03 | -0.04, -0.02 | 0.011 | 0.11 | 0.08, 0.15 | 5.87x10^-07^ |
|  | Former | -0.02 | -0.04, -0.01 | 0.032 | 0.09 | 0.05, 0.13 | 5.01x10^-05^ |
|  | Never | -5.49x10^-05^ | -0.01, 0.01 | 0.994 | -0.02 | -0.05, 0.01 | 0.131 |
| Getting up | Current | 0.02 | -0.00, 0.04 | 0.135 | -0.06 | -0.10, -0.02 | 0.004 |
|  | Ever | 0.01 | 0.00, 0.02 | 0.064 | -0.04 | -0.06, -0.02 | <0.001 |
|  | Former | 0.01 | -0.01, 0.02 | 0.371 | -0.04 | -0.06, -0.01 | 0.007 |
|  | Never | -0.00 | -0.01, 0.01 | 0.932 | 0.02 | -0.00, 0.04 | 0.069 |
| Insomnia symptoms | Current | -0.00 | -0.02, 0.02 | 0.879 | 0.02 | -0.01, 0.05 | 0.173 |
|  | Ever | 0.00 | -0.01, 0.01 | 0.608 | 0.00 | -0.02, 0.02 | 0.751 |
|  | Former | 0.00 | -0.01, 0.02 | 0.648 | -0.00 | -0.02, 0.02 | 0.890 |
|  | Never | -0.00 | -0.01, 0.01 | 0.955 | -0.02 | -0.03, 0.00 | 0.060 |
| Napping | Current | 0.04 | 0.02, 0.05 | 0.011 | 0.01 | -0.02, 0.04 | 0.608 |
|  | Ever | 0.00 | -0.00, 0.01 | 0.302 | 0.00 | -0.02, 0.02 | 0.831 |
|  | Former | -0.00 | -0.01, 0.01 | 0.606 | 0.00 | -0.02, 0.02 | 0.867 |
|  | Never | 0.00 | -0.00, 0.01 | 0.655 | 0.00 | -0.01, 0.02 | 0.630 |
| Daytime sleepiness | Current | 0.01 | -0.01, 0.02 | 0.360 | -0.07 | -0.01, -0.05 | 1.78x10^-06^ |
|  | Ever | 0.00 | -0.00, 0.01 | 0.592 | -0.02 | -0.03, -0.01 | 0.001 |
|  | Former | 0.00 | -0.01, 0.01 | 0.818 | -0.01 | -0.02, 0.01 | 0.328 |
|  | Never | -0.00 | -0.01, 0.00 | 0.237 | -0.01 | -0.02, 0.01 | 0.331 |
| Sleep duration | Current | -0.02 | -0.06, 0.02 | 0.388 | -0.01 | -0.07, 0.05 | 0.793 |
|  | Ever | -0.01 | -0.02, 0.01 | 0.351 | 0.04 | 0.01, 0.07 | 0.013 |
|  | Former | -0.00 | -0.02, 0.01 | 0.600 | 0.06 | 0.02, 0.09 | <0.001 |
|  | Never | 0.01 | -0.01, 0.02 | 0.384 | -0.02 | -0.05, 0.01 | 0.236 |

*Coded towards being an evening person

Supplementary Table S5. Univariable Mendelian randomisation analysis (weighted median) of nicotine metabolite ratio (NMR), cigarettes per day (CPD) and continuous sleep outcomes measured in UK Biobank stratified by smoking status

| Sleep outcome | Smoking status | NMR |  |  | CPD |  | |  |
| --- | --- | --- | --- | --- | --- | --- | --- | --- |
|  |  | Effect | 95% CI | P-value | Effect | 95% CI | P-value | |
| Chronotype | Current | -0.04 | -0.05, -0.02 | 7.04x10^-06^ | 0.17 | 0.13, 0.21 | | 9.61x10^-12^ |
|  | Ever | -0.03 | -0.03, -0.02 | 4.70x10^-14^ | 0.10 | 0.08, 0.12 | | 9.85x10^-26^ |
|  | Former | -0.02 | -0.03, -0.01 | 55.3x10^-06^ | 0.07 | 0.05, 0.09 | | 1.41x10^-11^ |
|  | Never | 0.00 | -0.01, 0.01 | 0.817 | -0.03 | -0.05, -0.02 | | 4.11x10^-05^ |
| Getting up | Current | 0.02 | 0.00, 0.03 | 0.007 | -0.06 | -0.09, -0.03 | | 4.09x10^-05^ |
|  | Ever | 0.01 | 0.00, 0.01 | 0.018 | -0.03 | -0.05, -0.02 | | 1.28x10^-05^ |
|  | Former | -0.00 | -0.01, 0.01 | 0.866 | -0.02 | -0.04, -0.01 | | 0.002 |
|  | Never | -0.00 | -0.01, 0.00 | 0.322 | 0.02 | 0.01, 0.03 | | 0.000 |
| Insomnia symptoms | Current | -0.00 | -0.01, 0.01 | 0.461 | 0.02 | -0.00, 0.05 | | 0.089 |
|  | Ever | -0.00 | -0.01, 0.00 | 0.153 | 0.01 | -0.00, 0.02 | | 0.188 |
|  | Former | -0.00 | -0.01, 0.00 | 0.204 | 0.00 | -0.01, 0.02 | | 0.566 |
|  | Never | 0.00 | -0.00, 0.00 | 0.914 | -0.01 | -0.02, 0.00 | | 0.081 |
| Napping | Current | 0.03 | 0.02, 0.04 | 1.56x10^-09^ | 0.02 | -0.01, 0.04 | | 0.159 |
|  | Ever | 0.00 | -0.00, 0.01 | 0.174 | 0.01 | -0.01, 0.02 | | 0.326 |
|  | Former | -0.00 | -0.01, 0.00 | 0.435 | 0.00 | -0.01, 0.01 | | 0.943 |
|  | Never | 0.00 | -0.01, 0.00 | 0.837 | 0.00 | -0.01, 0.01 | | 0.656 |
| Daytime sleepiness | Current | 0.01 | 0.00, 0.02 | 0.011 | -0.06 | -0.08, -0.04 | | 7.75x10^-09^ |
|  | Ever | -0.00 | -0.00, 0.00 | 0.500 | -0.02 | -0.03, -0.01 | | 0.002 |
|  | Former | -0.00 | -0.01, 0.00 | 0.170 | -0.00 | -0.01, 0.01 | | 0.556 |
|  | Never | -0.00 | -0.01, -0.00 | 0.012 | -0.01 | -0.01, 0.00 | | 0.112 |
| Sleep duration | Current | -0.02 | -0.01, -0.00 | 0.024 | -0.01 | -0.06, 0.04 | | 0.657 |
|  | Ever | -0.01 | -0.01, 0.00 | 0.075 | 0.04 | 0.02, 0.06 | | 4.13x10^-05^ |
|  | Former | -0.00 | -0.01, 0.01 | 0.486 | 0.06 | 0.04, 0.08 | | 3.19x10^-07^ |
|  | Never | 0.01 | -0.00, 0.01 | 0.131 | -0.02 | -0.04, -0.00 | | 0.014 |

*Coded towards being an evening person

Supplementary Table S6. Univariable Mendelian randomisation analysis (weighted mode) of nicotine metabolite ratio (NMR), cigarettes per day (CPD) and continuous sleep outcomes measured in UK Biobank stratified by smoking status

| Sleep outcome | Smoking status | NMR |  |  | CPD |  | |  |
| --- | --- | --- | --- | --- | --- | --- | --- | --- |
|  |  | Effect | 95% CI | P-value | Effect | 95% CI | P-value | |
| Chronotype | Current | -0.03 | 0.00, 0.03 | 0.063 | 0.17 | 0.13, 0.21 | | 2.59x10^-17^ |
|  | Ever | -0.03 | 0.00, 0.01 | 0.068 | 0.09 | 0.08, 0.11 | | 8.65x10^-16^ |
|  | Former | -0.02 | -0.01, 0.01 | 0.859 | 0.07 | 0.05, 0.08 | | 9.88x10^-10^ |
|  | Never | 0.00 | -0.01, 0.00 | 0.424 | -0.03 | -0.05, -0.02 | | 5.34x10^-05^ |
| Getting up | Current | 0.02 | 0.00, 0.03 | 0.062 | -0.06 | -0.09, -0.03 | | 3.83x10^-05^ |
|  | Ever | 0.01 | 0.00, 0.01 | 0.068 | -0.03 | -0.04, -0.02 | | 9.77x10^-06^ |
|  | Former | -0.00 | -0.01, 0.01 | 0.859 | -0.02 | -0.04, -0.01 | | 0.001 |
|  | Never | -0.00 | -0.01, 0.00 | 0.424 | 0.02 | 0.01, 0.03 | | 0.000 |
| Insomnia symptoms | Current | -0.00 | -0.01, 0.01 | 0.567 | 0.02 | 0.00, 0.05 | | 0.051 |
|  | Ever | -0.00 | -0.01, 0.00 | 0.221 | 0.01 | -0.00, 0.02 | | 0.151 |
|  | Former | -0.00 | -0.01, 0.00 | 0.340 | 0.00 | -0.01, 0.02 | | 0.658 |
|  | Never | 0.00 | -0.00, 0.01 | 0.944 | -0.01 | -0.02, 0.00 | | 0.135 |
| Napping | Current | 0.03 | 0.02, 0.04 | 0.002 | 0.01 | -0.01, 0.03 | | 0.170 |
|  | Ever | 0.00 | -0.00, 0.01 | 0.177 | 0.01 | -0.00, 0.02 | | 0.172 |
|  | Former | -0.00 | -0.01, 0.00 | 0.490 | 0.00 | -0.01, 0.01 | | 0.670 |
|  | Never | 0.00 | -0.01, 0.00 | 0.823 | 0.01 | -0.00, 0.01 | | 0.231 |
| Daytime sleepiness | Current | 0.01 | 0.00, 0.02 | 0.058 | -0.06 | -0.08, -0.04 | | 1.13x10^-08^ |
|  | Ever | -0.00 | -0.00, 0.00 | 0.705 | -0.02 | -0.03, -0.01 | | 4.36x10^-05^ |
|  | Former | -0.00 | -0.01, 0.0 | 0.249 | -0.01 | -0.01, 0.00 | | 0.225 |
|  | Never | -0.00 | -0.01, -0.00 | 0.065 | -0,00 | -0.01, 0.00 | | 0.276 |
| Sleep duration | Current | -0.02 | -0.04, -0.00 | 0.081 | -0.01 | -0.05, 0.04 | | 0.795 |
|  | Ever | -0.01 | -0.02, 0.02 | 0.182 | 0.01 | 0.03, 0.07 | | 1.04x10^-05^ |
|  | Former | -0.00 | -0.01, 0.01 | 0.525 | 0.06 | 0.04, 0.08 | | 3.29x10^-07^ |
|  | Never | 0.01 | -0.00, 0.01 | 0.186 | -0.02 | -0.04, -0.01 | | 0.003 |

*Coded towards being an evening person

Supplementary Table S7. Heterogeneity statistics for NMR MR using sleep outcomes in UK Biobank stratified by smoking status

| Sleep outcome | Smoking status | IVW-MR | | | MR-Egger | | |
| --- | --- | --- | --- | --- | --- | --- | --- |
|  |  | Q | Q df | P-value | Q | Q df | P-value |
| Chronotype | Current | 4.36 | 5 | 0.499 | 4.22 | 4 | 0.377 |
|  | Ever | 5.71 | 5 | 0.336 | 4.69 | 4 | 0.321 |
|  | Former | 3.16 | 5 | 0.675 | 2.31 | 4 | 0.678 |
|  | Never | 5.83 | 5 | 0.324 | 5.80 | 4 | 0.324 |
| Getting up | Current | 1.19 | 5 | 0.946 | 1.17 | 4 | 0.884 |
|  | Ever | 2.41 | 5 | 0.790 | 0.60 | 4 | 0.963 |
|  | Former | 2.88 | 5 | 0.718 | 1.49 | 4 | 0.829 |
|  | Never | 6.75 | 5 | 0.240 | 6.71 | 4 | 0.151 |
| Insomnia symptoms | Current | 4.25 | 5 | 0.400 | 4.05 | 4 | 0.514 |
|  | Ever | 6.63 | 5 | 0.274 | 5.13 | 4 | 0.249 |
|  | Former | 12.29 | 5 | 0.031 | 11.17 | 4 | 0.025 |
|  | Never | 0.34 | 5 | 0.999 | 0.26 | 4 | 0.991 |
| Napping | Current | 7.40 | 5 | 0.192 | 4.05 | 4 | 0.399 |
|  | Ever | 1.10 | 5 | 0.954 | 0.84 | 4 | 0.934 |
|  | Former | 0.84 | 5 | 0.974 | 0.79 | 4 | 0.940 |
|  | Never | 1.33 | 5 | 0.932 | 0.95 | 4 | 0.918 |
| Daytime sleepiness | Current | 3.65 | 5 | 0.600 | 3.64 | 4 | 0.457 |
|  | Ever | 1.81 | 5 | 0.874 | 0.55 | 4 | 0.968 |
|  | Former | 2.14 | 5 | 0.830 | 0.62 | 4 | 0.961 |
|  | Never | 0.89 | 5 | 0.971 | 0.87 | 4 | 0.930 |
| Sleep duration | Current | 7.30 | 5 | 0.200 | 7.27 | 4 | 0.122 |
|  | Ever | 0.60 | 5 | 0.988 | 0.57 | 4 | 0.966 |
|  | Former | 1.93 | 5 | 0.859 | 1.84 | 4 | 0.766 |
|  | Never | 1.49 | 5 | 0.914 | 1.48 | 4 | 0.830 |

*Coded towards being an evening person

Supplementary Table S8. Heterogeneity statistics for CPD MR using sleep outcomes in UK Biobank stratified by smoking status

| Sleep outcome | Smoking status | IVW-MR | | | MR-Egger | | |
| --- | --- | --- | --- | --- | --- | --- | --- |
|  |  | Q | Q df | P-value | Q | Q df | P-value |
| Chronotype | Current | 93.86 | 50 | <0.001 | 81.59 | 49 | 0.002 |
|  | Ever | 226.57 | 50 | 2.57x10^-24^ | 188.12 | 49 | 3.52x10^-18^ |
|  | Former | 194.06 | 50 | 7.51x10^-19^ | 162.60 | 49 | 4.21x10^-14^ |
|  | Never | 150.33 | 50 | 5.64x10^-12^ | 143.08 | 49 | 3.80x10^-11^ |
| Getting up | Current | 66.10 | 50 | 0.054 | 65.35 | 49 | 0.059 |
|  | Ever | 119.96 | 50 | 1.11x10^-07^ | 109.80 | 49 | 1.48x10^-06^ |
|  | Former | 114.59 | 50 | 5.59x10^-07^ | 103.50 | 49 | 8.99x10^-06^ |
|  | Never | 117.30 | 50 | 2.49x10^-07^ | 117.17 | 49 | 1.64x10^-07^ |
| Insomnia symptoms | Current | 52.23 | 50 | 0.387 | 52.03 | 49 | 0.357 |
|  | Ever | 94.52 | 50 | <0.001 | 96.06 | 49 | 9.77x10^-05^ |
|  | Former | 86.54 | 50 | 0.001 | 83.81 | 49 | 0.001 |
|  | Never | 100.87 | 50 | 2.74x10^-05^ | 87.61 | 49 | <0.001 |
| Napping | Current | 74.44 | 50 | 0.014 | 71.85 | 49 | 0.018 |
|  | Ever | 125.48 | 50 | 2.04x10^-08^ | 122.15 | 49 | 3.53x10^-08^ |
|  | Former | 102.68 | 50 | 1.67x10^-05^ | 102.05 | 49 | 1.34x10^-05^ |
|  | Never | 111.49 | 50 | 1.38x10^-06^ | 110.58 | 49 | 1.18x10^-06^ |
| Daytime sleepiness | Current | 87.71 | 50 | <0.001 | 76.09 | 49 | 0.008 |
|  | Ever | 78.29 | 50 | 0.006 | 75.69 | 49 | 0.009 |
|  | Former | 59.15 | 50 | 0.176 | 59.12 | 49 | 0.152 |
|  | Never | 73.26 | 50 | 0.018 | 73.07 | 49 | 0.014 |
| Sleep duration | Current | 59.86 | 50 | 0.160 | 59.79 | 49 | 0.139 |
|  | Ever | 94.42 | 50 | <0.001 | 94.36 | 49 | <0.001 |
|  | Former | 77.60 | 50 | 0.007 | 77.59 | 49 | 0.006 |
|  | Never | 123.71 | 50 | 3.49x10^-08^ | 123.63 | 49 | 2.22x10^-08^ |

*Coded towards being an evening person

Supplementary Table S9. Non-zero intercept terms of MR-Egger regression analyses from univariable Mendelian randomisation analysis of nicotine metabolite ratio (NMR), cigarettes per day (CPD) and continuous sleep outcomes measured in UK Biobank stratified by smoking status

| Sleep outcome | Smoking status |  | NMR |  | | | CPD | |
| --- | --- | --- | --- | --- | --- | --- | --- | --- |
|  |  | Effect | P-value | | Effect | | | P-value |
| Chronotype | Current | -0.00 | 0.739 | | | -0.01 | | 0.009 |
|  | Ever | 0.00 | 0.404 | | | -0.00 | | 0.003 |
|  | Former | 0.00 | 0.410 | | | -0.00 | | 0.003 |
|  | Never | 0.00 | 0.902 | | | -0.00 | | 0.122 |
| Getting up | Current | -0.00 | 0.877 | | | 0.00 | | 0.272 |
|  | Ever | -0.00 | 0.249 | | | 0.00 | | 0.038 |
|  | Former | -0.00 | 0.303 | | | 0.00 | | 0.026 |
|  | Never | -0.00 | 0.904 | | | 0.00 | | 0.820 |
| Insomnia symptoms | Current | -0.00 | 0.677 | | | -0.00 | | 0.664 |
|  | Ever | -0.00 | 0.340 | | | 0.00 | | 0.376 |
|  | Former | -0.00 | 0.561 | | | 0.00 | | 0.212 |
|  | Never | 0.00 | 0.810 | | | 0.00 | | 0.009 |
| Napping | Current | -0.01 | 0.144 | | | 0.00 | | 0.190 |
|  | Ever | -0.00 | 0.634 | | | 0.00 | | 0.256 |
|  | Former | 0.00 | 0.830 | | | 0.00 | | 0.587 |
|  | Never | -0.00 | 0.571 | | | 0.00 | | 0.528 |
| Daytime sleepiness | Current | -0.00 | 0.905 | | | 0.00 | | 0.009 |
|  | Ever | -0.00 | 0.324 | | | 0.00 | | 0.200 |
|  | Former | -0.00 | 0.286 | | | -7.83x10^-05^ | | 0.869 |
|  | Never | 0.00 | 0.904 | | | 0.00 | | 0.722 |
| Sleep duration | Current | -0.00 | 0.907 | | | -0.00 | | 0.813 |
|  | Ever | 0.00 | 0.874 | | | -0.00 | | 0.867 |
|  | Former | 0.00 | 0.777 | | | -9.64x10^-05^ | | 0.936 |
|  | Never | -0.00 | 0.924 | | | 0.00 | | 0.860 |

*Coded towards being an evening person

Supplementary Table S10. Multivariable Mendelian randomisation analysis (IVW-MVMR) of nicotine metabolite ratio (NMR), cigarettes per day (CPD) and continuous sleep outcomes measured in UK Biobank stratified by smoking status

| Sleep outcome | Smoking status | NMR |  |  | CPD |  | |  | Q |
| --- | --- | --- | --- | --- | --- | --- | --- | --- | --- |
|  |  | Effect | 95% CI | P-value | Effect | 95% CI | P-value | |  |
| Chronotype | Current | -0.04 | -0.06, -0.02 | 0.002 | 0.07 | 0.01, 0.13 | | 0.035 | 73.97 |
|  | Ever | -0.03 | -0.04, -0.01 | 0.003 | 0.04 | -0.01, 0.09 | | 0.135 | 186.16 |
|  | Former | -0.02 | -0.04, 0.00 | 0.081 | 0.02 | -0.03, 0.07 | | 0.515 | 175.43 |
|  | Never | 0.01 | -0.00, 0.02 | 0.161 | -0.05 | -0.08, -0.01 | | 0.008 | 141.82 |
| Getting up | Current | 0.02 | 0.00, 0.03 | 0.015 | -0.03 | 0.07, 0.01 | | 0.184 | 53.18 |
|  | Ever | 0.01 | -0.00, 0.02 | 0.268 | -0.01 | -0.03, 0.02 | | 0.636 | 111.18 |
|  | Former | -0.00 | -0.01, 0.01 | 0.715 | 0.00 | 0.03, 0.03 | | 0.785 | 109.49 |
|  | Never | -0.00 | -0.01, 0.01 | 0.478 | 0.02 | -0.01, 0.04 | | 0.136 | 119.73 |
| Insomnia symptoms | Current | -0.01 | -0.02, 0.01 | 0.263 | 0.01 | -0.02, 0.05 | | 0.499 | 50.68 |
|  | Ever | -0.00 | -0.01, 0.00 | 0.401 | 0.02 | -0.00, 0.04 | | 0.061 | 88.76 |
|  | Former | -0.00 | -0.01, 0.01 | 0.729 | 0.02 | -0.00, 0.05 | | 0.066 | 86.85 |
|  | Never | -0.00 | -0.01, 0.01 | 0.065 | 0.02 | -0.00 0.04 | | 0.075 | 86.53 |
| Napping | Current | 0.02 | 0.00, 0.03 | 0.029 | 0.04 | 0.00, 0.07 | | 0.054 | 69.85 |
|  | Ever | 0.00 | -0.01, 0.01 | 0.738 | 0.01 | -0.01, 0.04 | | 0.211 | 115.92 |
|  | Former | -0.00 | -0.01, 0.01 | 0.878 | 0.00 | -0.02, 0.03 | | 0.666 | 94.24 |
|  | Never | -0.00 | -0.01, 0.01 | 0.873 | 0.01 | -0.01, 0.02 | | 0.497 | 108.62 |
| Daytime sleepiness | Current | 0.01 | -0.00, 0.02 | 0.075 | -0.03 | -0.06, -0.00 | | 0.052 | 58.43 |
|  | Ever | 0.00 | -0.00, 0.01 | 0.876 | -0.01 | -0.02, 0.00 | | 0.191 | 70.34 |
|  | Former | -0.00 | -0.01, 0.00 | 0.542 | -0.00 | -0.02, 0.01 | | 0.536 | 56.57 |
|  | Never | -0.00 | -0.01, 0.00 | 0.403 | -0.00 | -0.02, 0.01 | | 0.442 | 69.64 |
| Sleep duration | Current | -0.02 | -0.04, 0.01 | 0.174 | -0.04 | -0.11, 0.02 | | 0.222 | 56.61 |
|  | Ever | -0.01 | -0.02, 0.00 | 0.154 | 0.03 | -0.01, 0.06 | | 0.127 | 87.21 |
|  | Former | -0.01 | -0.02, 0.01 | 0.224 | 0.05 | 0.01, 0.09 | | 0.011 | 74.85 |
|  | Never | 0.01 | -0.00, 0.02 | 0.169 | -0.02 | -0.05, 0.01 | | 0.285 | 103.62 |

*Coded towards being an evening person

Supplementary Table S11. Multivariable Mendelian randomisation analysis (MVMR-Egger) of nicotine metabolite ratio (NMR), cigarettes per day (CPD) and continuous sleep outcomes measured in UK Biobank stratified by smoking status

| Sleep outcome | Smoking status | NMR | | | CPD | | |
| --- | --- | --- | --- | --- | --- | --- | --- |
|  |  | Effect | 95% CI | P-value | Effect | 95% CI | P-value |
| Chronotype | Current | -0.04 | -0.07, -0.02 | 0.001 | 0.14 | 0.04, 0.24 | 0.011 |
|  | Ever | -0.03 | -0.05, -0.01 | 0.001 | 0.10 | 0.03, 0.17 | 0.011 |
|  | Former | -0.02 | -0.04 -0.00 | 0.035 | 0.08 | 0.01, 0.16 | 0.036 |
|  | Never | 0.01 | -0.00, 0.02 | 0.227 | -0.02 | -0.08, 0.03 | 0.452 |
| Getting up | Current | 0.02 | 0.00, 0.04 | 0.016 | -0.04 | -0.11, 0.03 | 0.287 |
|  | Ever | 0.01 | -0.00, 0.02 | 0.212 | -0.02 | -0.07, 0.02 | 0.281 |
|  | Former | -0.00 | -0.01, 0.01 | 0.869 | -0.02 | -0.07, 0.03 | 0.395 |
|  | Never | -0.00 | -0.01, 0.01 | 0.525 | 0.01 | -0.03, 0.05 | 0.529 |
| Insomnia symptoms | Current | -0.01 | -0.02, 0.01 | 0.266 | 0.02 | -0.04, 0.07 | 0.603 |
|  | Ever | -0.00 | -0.01, 0.00 | 0.415 | 0.02 | -0.02, 0.06 | 0.256 |
|  | Former | -0.00 | -0.02, 0.06 | 0.746 | 0.02 | -0.02, 0.06 | 0.287 |
|  | Never | -0.00 | -0.01, 0.01 | 0.871 | -0.01 | -0.04, 0.03 | 0.740 |
| Napping | Current | 0.02 | 0.00, 0.03 | 0.028 | 0.03 | -0.03, 0.08 | 0.394 |
|  | Ever | 0.00 | -0.01, 0.01 | 0.657 | 0.00 | -0.03, 0.04 | 0.842 |
|  | Former | -0.00 | -0.01, 0.01 | 0.939 | -0.00 | -0.04, 0.03 | 0.908 |
|  | Never | -0.00 | -0.01, 0.01 | 0.941 | 2.87x10^-05^ | -0.03, 0.03 | 0.998 |
| Daytime sleepiness | Current | 0.01 | 0.00, 0.02 | 0.022 | -0.07 | -0.11, -0.03 | 0.001 |
|  | Ever | 0.00 | -0.00, 0.01 | 0.807 | -0.01 | -0.03, 0.01 | 0.207 |
|  | Former | -0.00 | -0.01, 0.00 | 0.474 | 0.00 | -0.02, 0.03 | 0.825 |
|  | Never | -0.00 | -0.01, 0.00 | 0.446 | -0.01 | -0.03, 0.01 | 0.450 |
| Sleep duration | Current | -0.02 | -0.04, 0.01 | 0.186 | -0.04 | -0.15, 0.07 | 0.446 |
|  | Ever | -0.01 | -0.02, 0.00 | 0.175 | 0.02 | -0.04, 0.08 | 0.448 |
|  | Former | -0.01 | -0.02, 0.01 | 0.253 | 0.04 | -0.02, 0.10 | 0.171 |
|  | Never | 0.01 | -0.00, 0.02 | 0.183 | -0.02 | -0.07, 0.04 | 0.554 |

*Coded towards being an evening person
